# A cerebrospinal fluid metabolic signature of recovery in normal pressure hydrocephalus

**DOI:** 10.64898/2026.03.29.26349660

**Authors:** Lisa J. Duan, Mei E. Tiemeyer, Owen P. Leary, Amanda Hasbrouck, Caleb P. Schultz, Rohan Meier, Shanzeh Sayied, Natalie Amaral-Nieves, Jeannette Riley Brook, Naama Kanarek, Saud Alushaini, Maria Guglielmo, Konstantina A. Svokos, Petra M. Klinge, Alexander Fleischmann, Maria Grazia Ruocco, Boryana Petrova

## Abstract

Understanding the biological determinants of recovery of neurological function remains a major challenge. Normal pressure hydrocephalus (NPH) is characterized by urinary incontinence, gait impairment and cognitive decline. Notably, cerebrospinal fluid (CSF) drainage can substantially improve symptoms, making NPH one of the few potentially reversible causes of dementia. However, postoperative benefit varies considerably between patients, and reliable preoperative predictors are lacking, leaving patients exposed to invasive intervention with uncertain likelihood of clinical benefit. We therefore asked whether preoperative CSF metabolic profiles capture biological states associated with neurological recovery.

We studied a single-center cohort of 86 patients with NPH (mean age 74.7 years, range 59–90; 44 men, 42 women). We analyzed ventricular CSF from patients undergoing shunt surgery by semi-targeted (n = 42) and targeted (n = 55) mass spectrometry, with additional lumbar CSF (n = 29) and plasma (n = 33). Pathways identified in the discovery cohort were summarized as indices and tested in the targeted cohort on a locked dataset with a prespecified statistical analysis plan.

A one-carbon/redox index, combining transsulfuration and choline metabolites, was associated with the number of symptom domains improved at 3 months (odds ratio 0.49 per SD, 95% CI 0.24–0.92; false discovery rate 0.051), most strongly for cognition, where lower index values accompanied improvement. Transcripts of the one-carbon pathway in CSF extracellular vesicles of the same patients shifted in the same direction (permutation p = 0.023; not significant after correction). Under nested cross-validation, a sparse metabolite model distinguished patients who improved cognitively, both in a leave-one-out implementation of our initial pipeline (AUC 0.79; permutation p = 0.028) and in the locked repeated 5-fold protocol (AUC 0.70; p = 0.046 after correction for model choice), and adding the prespecified indices to clinical covariates improved cognitive prediction; the prespecified full-metabolome model and models for gait and urinary outcomes did not exceed chance.

These results point to one-carbon and choline metabolism as a candidate biological correlate of cognitive recovery after shunt surgery, to be tested in a prospective validation study.

## Introduction

The biological states associated with neurological recovery remain poorly understood, particularly in neurological disorders where therapeutic responses are heterogeneous and recovery is often limited. Normal pressure hydrocephalus (NPH) is a progressive disorder of cerebrospinal fluid (CSF) dynamics in older adults, characterized by ventriculomegaly together with urinary incontinence, gait disturbance and cognitive impairment.^1–4^ NPH is clinically progressive, affects approximately 20 million people worldwide and accounts for ∼5% of dementia cases.^5–9^ Among the few potentially reversible causes of dementia, NPH is often treatable with CSF drainage via ventriculoperitoneal shunt (VPS) surgery, which can result in substantial recovery of mobility, cognition, and quality of life.^10–12^ However, clinical responses vary, and not all patients demonstrate sustained functional benefit across all symptom domains.^2,4,10,13–17^ This uncertainty presents a challenge for patient selection: surgery may be unnecessary in some patients and exposes others to procedural risks, whereas deferring surgery may withhold a potentially effective treatment. Clinical management is further complicated by symptom overlap and frequent comorbidity with other neurodegenerative disorders, including Alzheimer’s and Parkinson’s diseases,^2,18–20^ as well as other age-related comorbidities.^21^ Together, these features suggest that treatment responsiveness in NPH may reflect underlying biological states not captured by current clinical assessment.

Preoperative patient stratification currently relies primarily on short-term symptomatic improvement following CSF drainage, most commonly via a high-volume lumbar puncture (HVLP), or “lumbar tap test”. Patients who show clinical improvement after HVLP are generally considered likely to benefit from shunt surgery, whereas lack of improvement does not reliably exclude treatment response. Consistently, HVLP has a high positive predictive value (∼90%) but a low negative predictive value (20–50%) as many patients who fail to improve during testing may still benefit from surgery.^3,22–28^ Consequently, clinical decision-making remains uncertain: a lack of confirmatory tests exposes some patients to unnecessary surgical risk, while over-reliance on HVLP likely leads to missed therapeutic opportunities for others. These limitations highlight the need for molecular biomarkers that capture recovery potential and complement current diagnostic approaches, improving patient stratification and clinical decision-making.

Emerging approaches include advanced imaging biomarkers,^29,30^ including machine-learning analysis of cortical thickness,^31^ EEG-based methods,^32,33^ and fluid dynamic monitoring techniques^34^; however, these strategies have yet to provide robust and clinically actionable predictors of treatment response.

CSF provides a particularly informative substrate for investigating neurological diseases because it directly reflects processes occurring within the central nervous system. Established CSF markers of neurodegeneration, including neurofilament light chain (NfL), t-tau, and AD-related amyloid/tau signatures, particularly reduced Aβ42 and composite ratios such as p-tau/Aβ42 or Aβ42/40, have been associated with shunt outcome, but with limited and inconsistent predictive accuracy^35–39^; most recently, CSF p-tau181 was not associated with shunt response in 230 shunted patients across two assay platforms.^40^ More recent proteomic studies have identified inflammatory markers associated with shunt surgery outcomes,^41,42^ while transcriptomic profiling of CSF-derived extracellular vesicles has revealed genes linked to postoperative improvement.^43^ Together, these findings suggest that molecular signatures of treatment responsiveness exist, although their translation into consistent and clinically applicable predictive frameworks remains limited.^44^

Metabolomics captures an orthogonal layer of molecular information by measuring small-molecule pathway activity and the physiological state of tissues. Because metabolites represent downstream products of cellular signaling and metabolic networks integrating genetic, environmental, and lifestyle influences, metabolomic profiling provides a functional readout of dynamic biological state. Recovery after shunt surgery in NPH may depend not only on the extent of neurological damage but also on the capacity of brain tissue to restore metabolic homeostasis and neuronal function; because the CSF metabolome reflects this capacity, it may carry information about recovery that clinical and structural measures do not.

At the same time, clinical metabolomics studies are frequently challenged by variability in analytical platforms, cohort sizes, and study design, complicating cross-study reproducibility and biomarker validation.^45^ While metabolomics investigations in NPH have begun to characterize disease-associated alterations (most recently, a targeted CSF panel that improved discrimination of iNPH from Alzheimer’s disease, mild cognitive impairment, and frontotemporal dementia^46^), their utility for predicting VPS outcomes has not been systematically explored. We therefore asked whether preoperative CSF metabolomic profiles capture biological states associated with recovery potential and can differentiate patients with divergent treatment outcomes.

To address this question, we developed a metabolomics-centered framework combining biomarker discovery, targeted analysis, multiple biofluids, and independent analytical platforms. Analyses were anchored in ventricular CSF to capture central nervous system biology most proximal to the primary pathology, with additional profiling of lumbar CSF and plasma to assess the extent to which these molecular signatures are reflected in clinically accessible biofluids. Because small cohorts and many candidate metabolites make outcome associations easy to overstate, pathway hypotheses from the discovery stage were summarized as indices and tested on a locked dataset with a statistical analysis plan fixed before the formal outcome analysis, and predictive performance was estimated by nested cross-validation against a permutation null. We then examined the biological context of the resulting signals in a separate transcriptomic dataset from the same patients.

## Materials and methods

### Patients and clinical outcomes

Patients evaluated for suspected normal pressure hydrocephalus (NPH) at Rhode Island Hospital/Brown University Health between January 2020 and April 2025 gave written informed consent to research sample donation during routine procedures. The study was approved by the Institutional Review Board of Rhode Island Hospital (IRB 1492994). The study was not registered, and patients and the public were not involved in its design, conduct, reporting or dissemination. Of 129 consenting patients, 86 were diagnosed with NPH (82 idiopathic, 4 secondary), did not withdraw and had at least one research sample; 80 of them underwent shunt surgery (Fig. 1B; Table 1). Ventricular CSF was available from 67 patients, enrolled in two consecutive cohorts: cohort 1 (earlier enrollment, n = 42) and cohort 2 (later enrollment, n = 25); the other 19 patients contributed lumbar CSF or plasma only. A diagnostic high-volume lumbar puncture (HVLP) was part of the routine clinical work-up in 48 patients (Table 1); its results were not available for this analysis.

**Figure 1.**
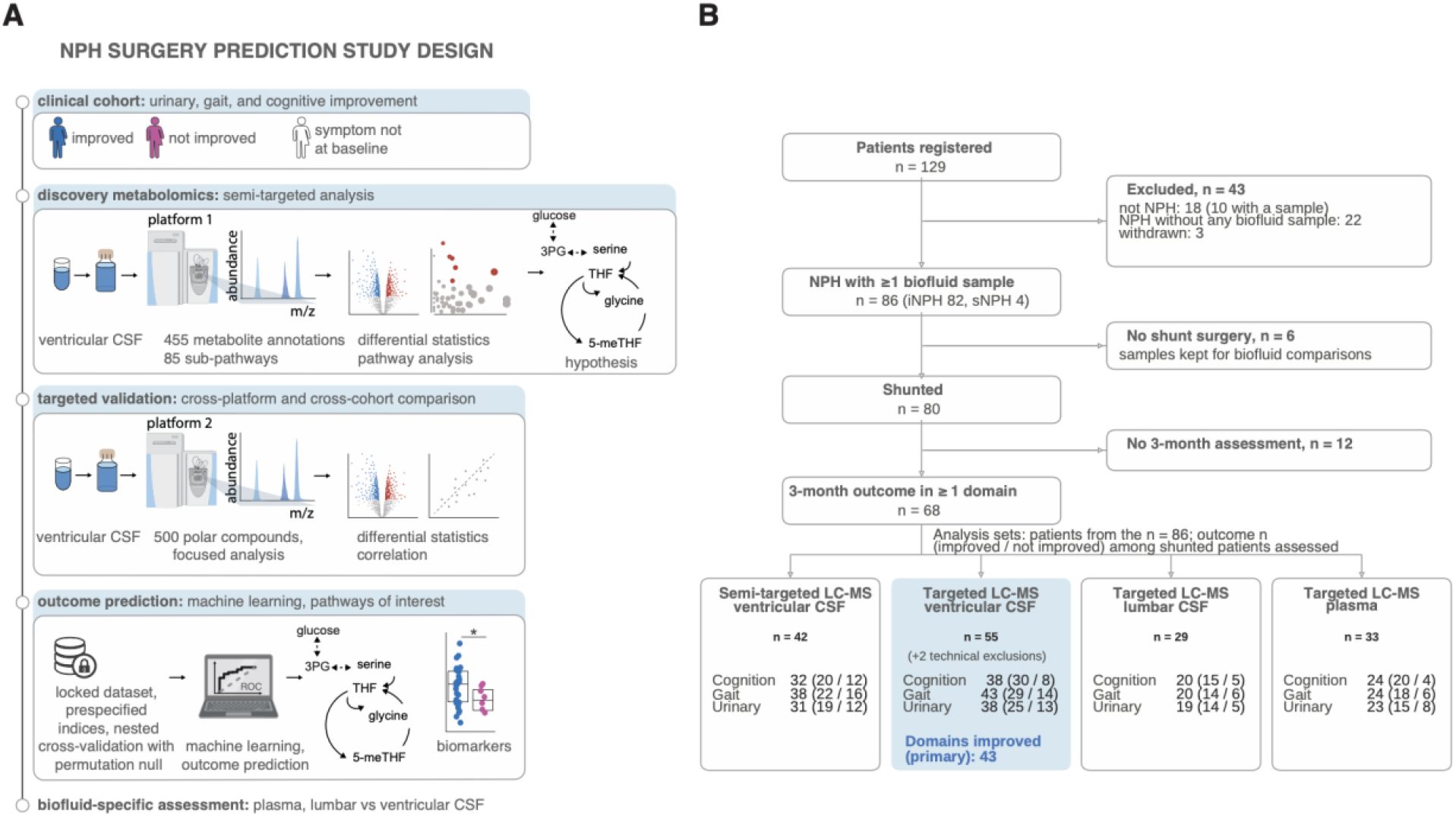
Study design and analytical framework. (**A**) Schematic of the two-stage design. Clinical cohort: urinary, gait and cognitive improvement at 3 months were scored per domain (improved, blue; not improved, magenta; symptom absent at baseline, white). Discovery metabolomics: ventricular CSF of cohort 1 was profiled on a semi-targeted LC–MS platform (platform 1; >1,000 features, 455 metabolite annotations, 9 super-pathways) and analyzed by differential statistics and pathway analysis to generate hypotheses. Targeted validation: ventricular CSF of cohorts 1 and 2 was profiled on a targeted LC–MS platform (platform 2; ∼500 polar compounds in the library, 155 metabolites passing quality control) for cross-platform and cross-cohort comparison. Outcome prediction: on a locked dataset, prespecified pathway indices were tested and prediction was evaluated by nested cross-validation with a permutation null. Biofluid-specific assessment: matched lumbar CSF and plasma were compared with ventricular CSF. (**B**) Patient flow schematic. Of 129 patients registered with suspected NPH, 43 were excluded: 18 not NPH (10 of whom had a sample), 22 NPH without any biofluid sample, and 3 withdrawn. This left 86 NPH patients with at least one biofluid sample (iNPH 82, sNPH 4). Six did not undergo shunt surgery; their samples were kept for biofluid comparisons. Of the 80 shunted patients, 12 had no 3-month assessment, leaving 68 with a 3-month outcome in at least one domain. Analysis sets give the number of patients with each outcome (improved / not improved) among shunted patients assessed: semi-targeted LC–MS, ventricular CSF (n = 42): cognition 32 (20 / 12), gait 38 (22 / 16), urinary 31 (19 / 12 targeted LC–MS, ventricular CSF (n = 55): cognition 38 (30 / 8), gait 43 (29 / 14), urinary 38 (25 / 13); number of domains improved (primary outcome), n = 43 targeted LC–MS, lumbar CSF (n = 29): cognition 20 (15 / 5), gait 20 (14 / 6), urinary 19 (14 / 5) targeted LC–MS, plasma (n = 33): cognition 24 (20 / 4), gait 24 (18 / 6), urinary 23 (15 / 8) Two targeted ventricular samples were excluded for technical reasons (one not acquired, one failed predefined quality control). No sample was excluded on the basis of its principal component analysis position or outcome. *Alt text:* Two-part figure. A: schematic of the study, flowing from the clinical cohort, shown as patient icons coloured by outcome in each symptom domain, through semi-targeted discovery and targeted validation in ventricular CSF, to outcome prediction and to comparison with lumbar CSF and plasma. B: patient flow diagram from 129 registered patients to 86 with NPH and at least one sample, 80 who underwent shunt surgery and 68 with a 3-month outcome, with boxes giving the number of patients in each analysis set and outcome.

**Table 1.**
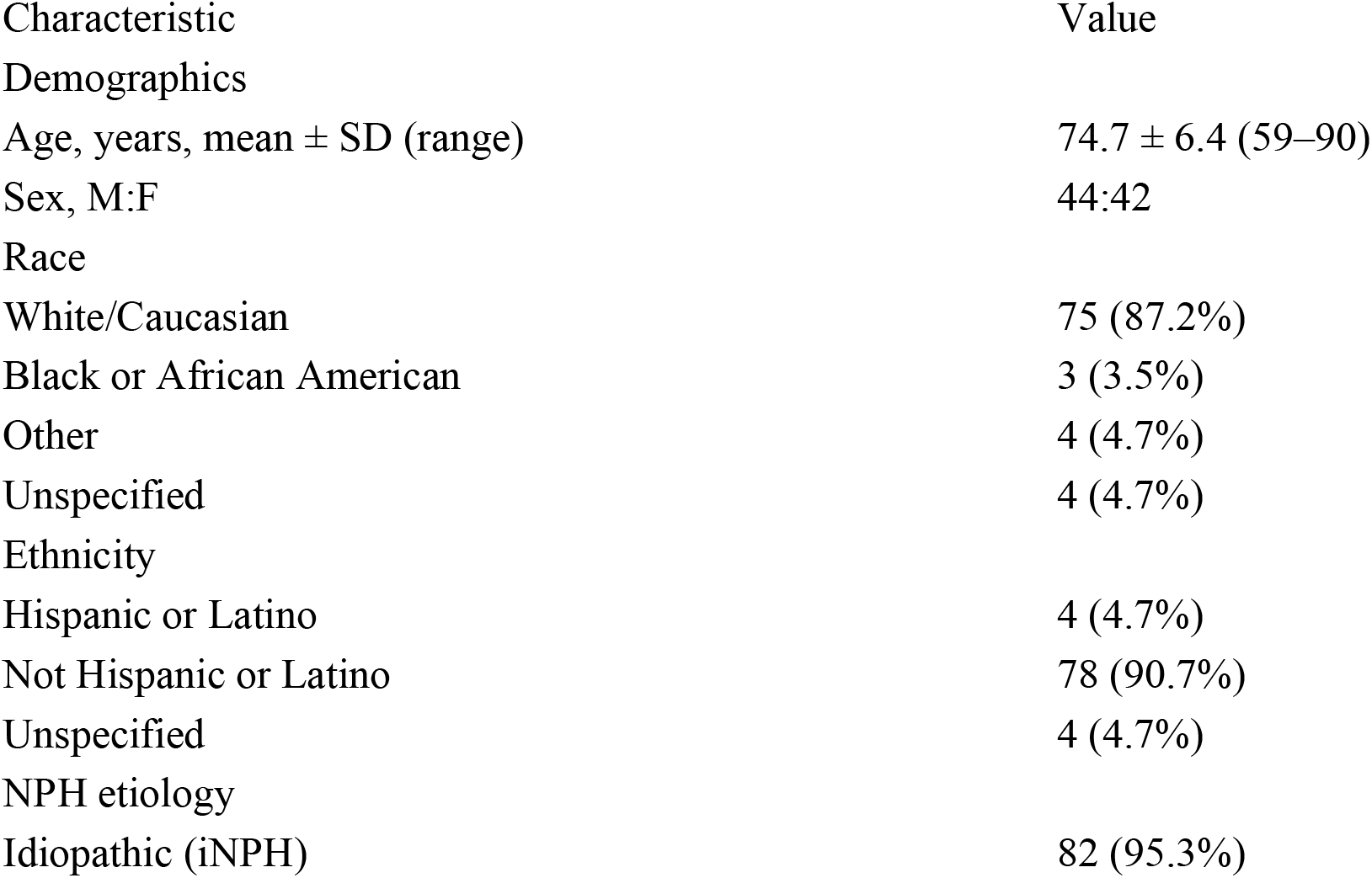
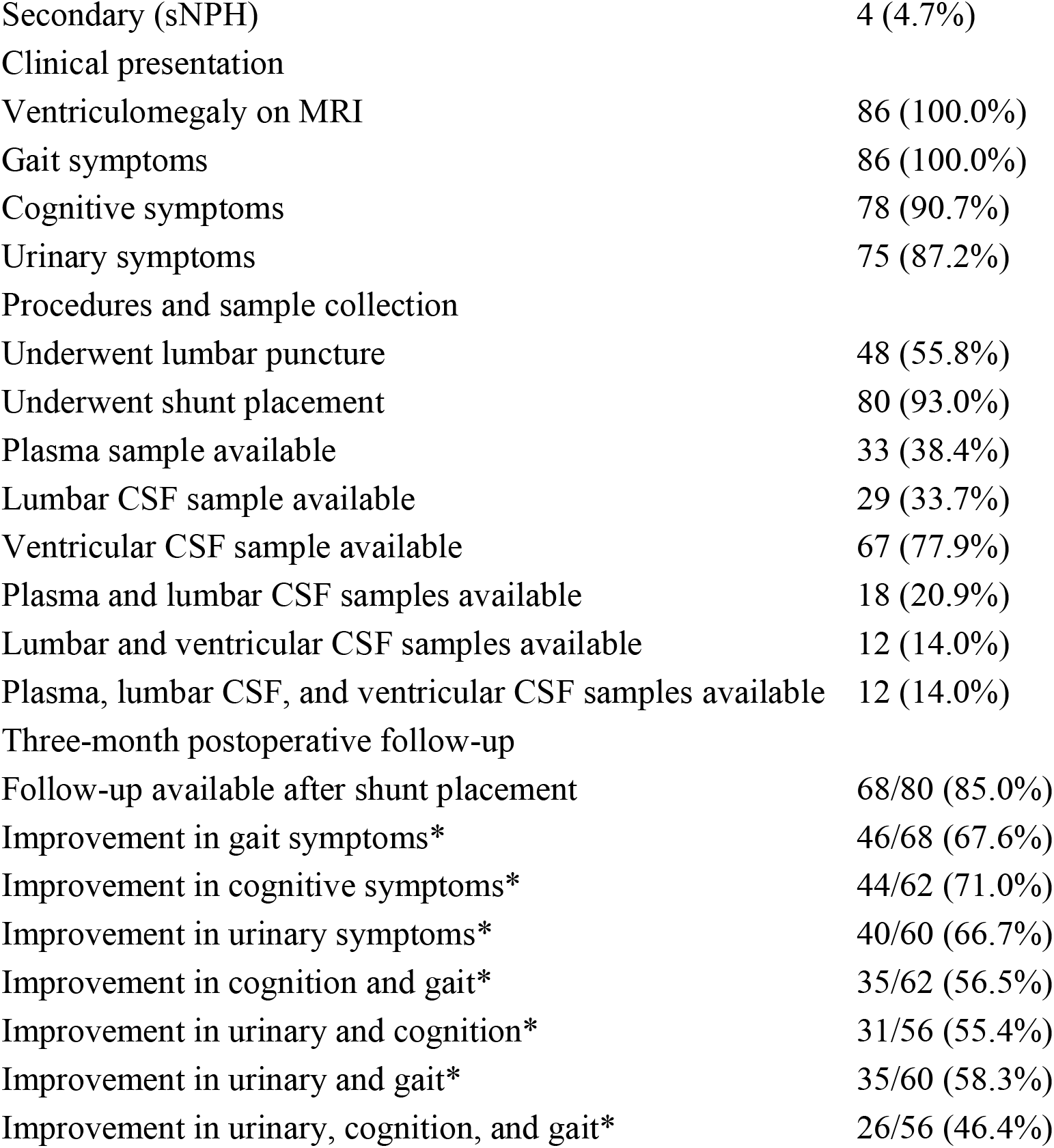
Demographic characteristics, clinical presentation, procedures, and postoperative outcomes of the study cohort (n = 86). Data are presented as n (%) unless otherwise indicated. Percentages for postoperative outcomes were calculated among patients with the corresponding symptom present at baseline and available 3-month follow-up data. CSF, cerebrospinal fluid; MRI, magnetic resonance imaging; NPH, normal pressure hydrocephalus; iNPH, idiopathic NPH; sNPH, secondary NPH.

| Characteristic | Value |
|---|---|
| Demographics |  |
| Age, years, mean ± SD (range) | 74.7 ± 6.4 (59–90) |
| Sex, M:F | 44:42 |
| Race |  |
| White/Caucasian | 75 (87.2%) |
| Black or African American | 3 (3.5%) |
| Other | 4 (4.7%) |
| Unspecified | 4 (4.7%) |
| Ethnicity |  |
| Hispanic or Latino | 4 (4.7%) |
| Not Hispanic or Latino | 78 (90.7%) |
| Unspecified | 4 (4.7%) |
| NPH etiology |  |
| Idiopathic (iNPH) | 82 (95.3%) |
| Secondary (sNPH) | 4 (4.7%) |
| Clinical presentation |  |
| Ventriculomegaly on MRI | 86 (100.0%) |
| Gait symptoms | 86 (100.0%) |
| Cognitive symptoms | 78 (90.7%) |
| Urinary symptoms | 75 (87.2%) |
| Procedures and sample collection |  |
| Underwent lumbar puncture | 48 (55.8%) |
| Underwent shunt placement | 80 (93.0%) |
| Plasma sample available | 33 (38.4%) |
| Lumbar CSF sample available | 29 (33.7%) |
| Ventricular CSF sample available | 67 (77.9%) |
| Plasma and lumbar CSF samples available | 18 (20.9%) |
| Lumbar and ventricular CSF samples available | 12 (14.0%) |
| Plasma, lumbar CSF, and ventricular CSF samples available | 12 (14.0%) |
| Three-month postoperative follow-up |  |
| Follow-up available after shunt placement | 68/80 (85.0%) |
| Improvement in gait symptoms* | 46/68 (67.6%) |
| Improvement in cognitive symptoms* | 44/62 (71.0%) |
| Improvement in urinary symptoms* | 40/60 (66.7%) |
| Improvement in cognition and gait* | 35/62 (56.5%) |
| Improvement in urinary and cognition* | 31/56 (55.4%) |
| Improvement in urinary and gait* | 35/60 (58.3%) |
| Improvement in urinary, cognition, and gait* | 26/56 (46.4%) |

Clinical assessment followed a previously described protocol.^43^ Urinary, gait and cognitive symptoms were recorded as present or absent at baseline. Each symptomatic domain was classified as improved or not improved at 3 months by comparing post-operative with pre-operative clinical documentation.^13^ Outcomes were assessed by the treating neurosurgeons and abstracted by study staff blinded to the metabolomic data. The criteria for improvement in each domain are given in the Supplementary Methods. Worsening was pooled with no improvement and kept as a separate flag. The primary outcome was the number of domains improved (0–3) among domains symptomatic at baseline.

### Biofluid sampling and metabolomics

Ventricular CSF was collected at catheter placement during shunt surgery, lumbar CSF at diagnostic lumbar puncture, and plasma (EDTA) by venipuncture. All samples were flash-frozen and stored at −80 °C (details in Supplementary Methods). Ventricular CSF from cohort 1 was profiled on a semi-targeted UHPLC–MS platform (Precision Metabolomics, Metabolon Inc., Durham, NC, USA). Spectra were matched against a library of more than 3,300 purified standards, and 455 metabolite annotations, including annotated degradation products, spanning 9 super-pathways and 85 sub-pathways were retained after quality control (Supplementary Data 1). Ventricular CSF from both cohorts, lumbar CSF and plasma were profiled on a targeted hydrophilic interaction LC–MS platform (ZIC-pHILIC, Q Exactive Orbitrap; Thermo Fisher Scientific). Annotation used an in-house retention-time library of about 500 authenticated compounds and 37 isotopically labelled internal standards (Supplementary Data 2).

Metabolites were retained if their peak area was linear across a dilution series of pooled quality-control samples (R² ≥ 0.8) and their coefficient of variation was below 30%. On both platforms, values were normalized to internal standards and to total biological material.^47^ On the targeted platform, 156 metabolite columns passed quality control; two of them are the same peak (N-formylmethionine) integrated twice, giving 155 distinct metabolites, and both columns were kept in the locked analyses. Two targeted ventricular samples were excluded for technical reasons only, by a rule fixed in advance: one was not acquired and one failed quality control. No sample was excluded on the basis of its metabolite profile. This left 42 semi-targeted ventricular, 55 targeted ventricular (31 from cohort 1, 24 from cohort 2), 29 lumbar and 33 plasma samples.

### Locked dataset and prespecified indices

Before any association with outcome was examined, the dataset was locked and the statistical analysis plan (indices, models, covariates and multiple-testing families) was fixed on 12 October 2026. Two composite indices and eleven ratios summarized the pathways identified in the semi-targeted discovery analysis. Their component metabolites were also informed by an initial exploratory analysis of the targeted data, in which metabolites, pathways and leave-one-out classifiers were selected and evaluated on the same patients (Supplementary Table 3d). The index tests were fixed before the reported analysis but are not independent of these data and require external validation.

To capture coordinated metabolic processes rather than individual metabolite changes, we constructed two pathway-based indices reflecting one-carbon/redox metabolism (P1) and tryptophan–kynurenine metabolism (P2). The one-carbon/redox index (P1) is the mean of signed z-scores of log₂ cystathionine (+), 2-hydroxybutyrate (−), 2-aminobutyrate (−), 5-methyltetrahydrofolate (+), phosphocholine (+) and phosphoethanolamine (+). Component signs were fixed from pathway position. The kynurenine index (P2) combines tryptophan, kynurenine and their ratio. The eleven ratios and the rules for missing values are defined in the Supplementary Methods. All indices were computed blind to outcome. Every later change to the dataset or analyses is dated in an analysis log released with the code.

### Statistical analysis

#### Prespecified index analyses

The prespecified indices (P1, P2 and the eleven ratios) were tested per SD. The primary analysis regressed the number of domains improved on each index by proportional-odds regression, adjusted for age, sex and *enrollment* cohort (n = 43; likelihood-ratio p-values, profile-likelihood 95% CIs.^48^ Per-domain logistic models with the same covariates were exploratory and used Firth’s penalized likelihood^49^ (Supplementary Methods). Likelihood-ratio tests replaced the Wald tests of the analysis plan after review by the study statistician; Wald results are given in the released code. Sensitivity analyses excluded patients who worsened, restricted the sample to idiopathic NPH, to cohort 2 or to patients with all three domains scored, excluded samples with flagged normalization factors or index values with |robust z| > 3.5, added body mass index, and used a quasi-binomial model of the proportion of domains improved (Supplementary Table 1b). Odds ratios above 1 indicate that higher index values accompany more improvement.

#### Multiple testing

P1 and P2 formed the primary family and the eleven ratios the secondary family. False discovery rate (FDR) was controlled within each family by the Benjamini–Hochberg procedure.^50^ In an exploratory per-metabolite scan, every endogenous targeted metabolite detected in at least half of the samples (n = 129) was tested with the same models, with Benjamini–Hochberg correction within each outcome. The same models gave the fold changes and significance shown on the pathway maps. Fold changes are expressed as improved over not improved throughout.

#### Outcome prediction

nested cross-validation and permutation testing. Two nested strategies were used (Fig. 4A; Supplementary Fig. 4A). First, the leave-one-out pipeline of our initial exploratory analysis was re-run after the lock in nested form for all three outcomes: within each leave-one-out training fold, log2-transformed metabolite values were autoscaled (as in the initial analysis) and ranked by an L2-penalized logistic regression and a linear support vector machine (SVM) was fitted on the top-k metabolites, with k and C chosen by an inner stratified 5-fold cross-validation (class-balanced weights; 10 random seeds for the inner split). Second, prediction followed a locked protocol for all outcomes, modelled from the targeted metabolome of the 55 ventricular samples (cognition n = 38, gait n = 43, urinary n = 38). Performance was estimated by nested cross-validation.^51^ The outer loop was stratified 5-fold cross-validation repeated 20 times, with folds identical for every model. An inner stratified 5-fold loop handled all tuning, feature selection and preprocessing, so test folds were never seen during model building. The prespecified primary model was ridge-penalized logistic regression on all metabolites. The comparators were a linear SVM and a nonlinear SVM (RBF kernel), each trained on the top-ranked metabolites selected within the training fold. In both strategies, significance was assessed by permuting outcome labels 1,000 times and rerunning the whole nested pipeline; in the locked protocol, with a max-over-models correction for the choice among the three models. Incremental value was assessed on the same folds by comparing log-loss between ridge models with clinical covariates (age, sex, cohort) alone, with P1 and P2 added, and with the full metabolome added. The non-nested estimates of the initial analysis and a permutation null of that procedure are given in Supplementary Table 3d.

#### Discovery and secondary analyses

For discovery, semi-targeted and targeted data were analyzed in MetaboAnalyst^52^ (differential statistics with Benjamini–Hochberg FDR, KEGG pathway analysis^53^ and PLS-DA). CSF extracellular-vesicle transcriptomes from cohort 143 were integrated by enzyme–metabolite correlations fixed before the transcriptomic data were opened and by competitive gene-set tests.^54^ Agreement between ventricular and lumbar CSF was assessed blind to outcome in 12 matched patients (correlation, Bland–Altman analysis^55^ and equivalence within ±1.5-fold.^56^ Full specifications of all analyses are given in the Supplementary Methods.

#### Software and availability

Analyses used Python 3.13 (NumPy, SciPy, scikit-learn 1.9.1^57^; regression models are implemented in the released code), R (cross-platform correlation), MetaboAnalyst 5.0–6.0 and GraphPad Prism 11. Code, the analysis log and file hashes are publicly available (see Data availability); patient-level data can be made available upon request. No final prediction model is released for use, because the prediction analyses were exploratory and did not provide reliable individual-level predictions.

## Results

### Study design for profiling recovery-associated metabolic states in NPH

To identify metabolic signatures associated with shunt responsiveness, we performed a two-stage metabolomics study comprising a discovery screen (semi-targeted) followed by targeted evaluation (Fig 1A and B). CSF was collected intraoperatively (ventricular CSF) during VPS surgery and, in a subset of patients, prospectively collected via lumbar puncture (lumbar CSF), providing direct access to central nervous system metabolic profiles. Plasma samples were also available for some patients, enabling assessment of biofluid-specific correlations across CSF compartments and peripheral blood (Fig. 1B; Supplementary Fig. 1A–C). Clinical phenotyping and outcome assessment followed routine protocols.^43^ For patients who underwent surgery, the routine postoperative clinical evaluation at 3 months was used for primary endpoint collection in outcomes-focused analyses,^4^ as further summarized in the Methods and **Table** 1.

A convenience sample of 129 patients presenting to neurosurgical clinics at a single center with suspected NPH were recruited between 1/2020 and 4/2025. Of those enrolled, 18 were not ultimately diagnosed with NPH based on clinical examinations, 3 withdrew from the study after enrolling, and of those remaining, no research samples were collected in 22 cases. At least one research sample (lumbar CSF, preoperative plasma, and/or ventricular CSF) was collected and analyzed from a total of n=86 NPH patients, comprising the final analytic cohort (summarized in **Table 1** and **Fig. 1B and Supplementary Fig. 1A**).

The mean age of the analytic cohort was 74.7±6.4 years (range 59–90), and 51.2% were male. 82 patients were diagnosed with idiopathic NPH (iNPH, 95.3%) and four were diagnosed with secondary NPH (sNPH, 4.7%). All patients presented with ventriculomegaly on Magnetic Resonance Imaging (MRI). In terms of the symptoms of the NPH diagnostic triad, urinary dysfunction was present in 87.2%, gait disturbance in 100.0%, and cognitive impairment in 90.7%. Among the analytic cohort, 48 (55.8%) underwent diagnostic HVLP and 80 (93.0%) underwent VPS placement. A total of 67 ventricular CSF samples, 29 lumbar CSF samples, and 33 preoperative plasma samples were obtained.

Of the patients who underwent VPS placement, 68 (85.0% of 80) completed the 3-month follow-up. Clinical improvement was assessed independently across urinary, gait, and cognitive domains. Improvement rates (calculated out of those who had each symptom present at baseline) were 66.7%, 67.6%, and 71.0%, for urinary, gait, and cognitive domains, respectively. Gait improvement was consistent with previously reported rates,^26,58,59^ whereas urinary and cognitive improvements exceeded those reported in prior studies.^3,58,60^ Gait and urinary responses showed partial co-occurrence, whereas cognitive improvement was less coupled to the other outcomes (**Supplementary Fig. 1E**). We therefore defined, in the analysis plan, the number of domains improved (0–3, among domains symptomatic at baseline) as the primary outcome, which uses all three domains in one ordinal scale, and analyzed each domain separately as a secondary, exploratory outcome. Patients who worsened in a domain (n = 5 in the primary analysis set) were analyzed together with those who did not improve.

Patients who underwent VPS surgery and returned for 3-month postoperative visits (n=68) were included in analyses of outcomes. In an initial biomarker discovery phase, 42 ventricular CSF samples (cohort 1) were profiled by semi-targeted metabolomics to identify candidate metabolites and pathways (**Supplementary Fig. 1B**; cognition n = 32, gait n = 38, urinary n = 31). These findings were subsequently evaluated using a targeted assay in an expanded set of 55 ventricular CSF samples that passed quality control, comprising 31 cohort 1 patients also profiled by the semi-targeted platform and 24 newly collected patients (cohort 2) (**Supplementary Fig. 1C**; cognition n = 38, gait n = 43, urinary n = 38). Two targeted samples were excluded for technical reasons only (one not acquired, one failed predefined quality-control criteria; Methods); no sample was excluded on the basis of its metabolite profile or outcome. For biofluid-specific assessment, 29 lumbar CSF and 33 plasma samples were analyzed on the targeted platform and compared against the targeted ventricular CSF cohort, irrespective of follow-up availability.

Across platforms, metabolite measurements demonstrated consistent cross-platform behavior, with 50% of metabolites showing both high correlation and quantitative agreement, and a smaller subset (7%) exhibiting strong correlation but systematic differences in absolute quantification (**Supplementary Fig. 1F)**. This partially overlapping two-stage design enabled biomarker discovery and hypothesis generation, followed by a test of the resulting hypotheses in an expanded cohort, while reducing cohort- and platform-specific effects commonly encountered in clinical metabolomics.^45,61,62^

### Semi-targeted metabolomics identifies metabolomic pathways associated with improvement in urinary, gait, and cognitive symptoms after shunt surgery

Semi-targeted LC–MS screening quantified 455 named metabolites in ventricular CSF, spanning 9 super-pathways (including amino acids, lipids, nucleotides and xenobiotics) and 85 sub-pathways, which were carried forward for downstream analyses (**Fig. 2A**; **Supplementary Data 1**). Quality assessment demonstrated high data consistency, with no outlier samples detected and no major differences observed between female and male patients (**Supplementary Fig. 2A**).

**Figure 2.**
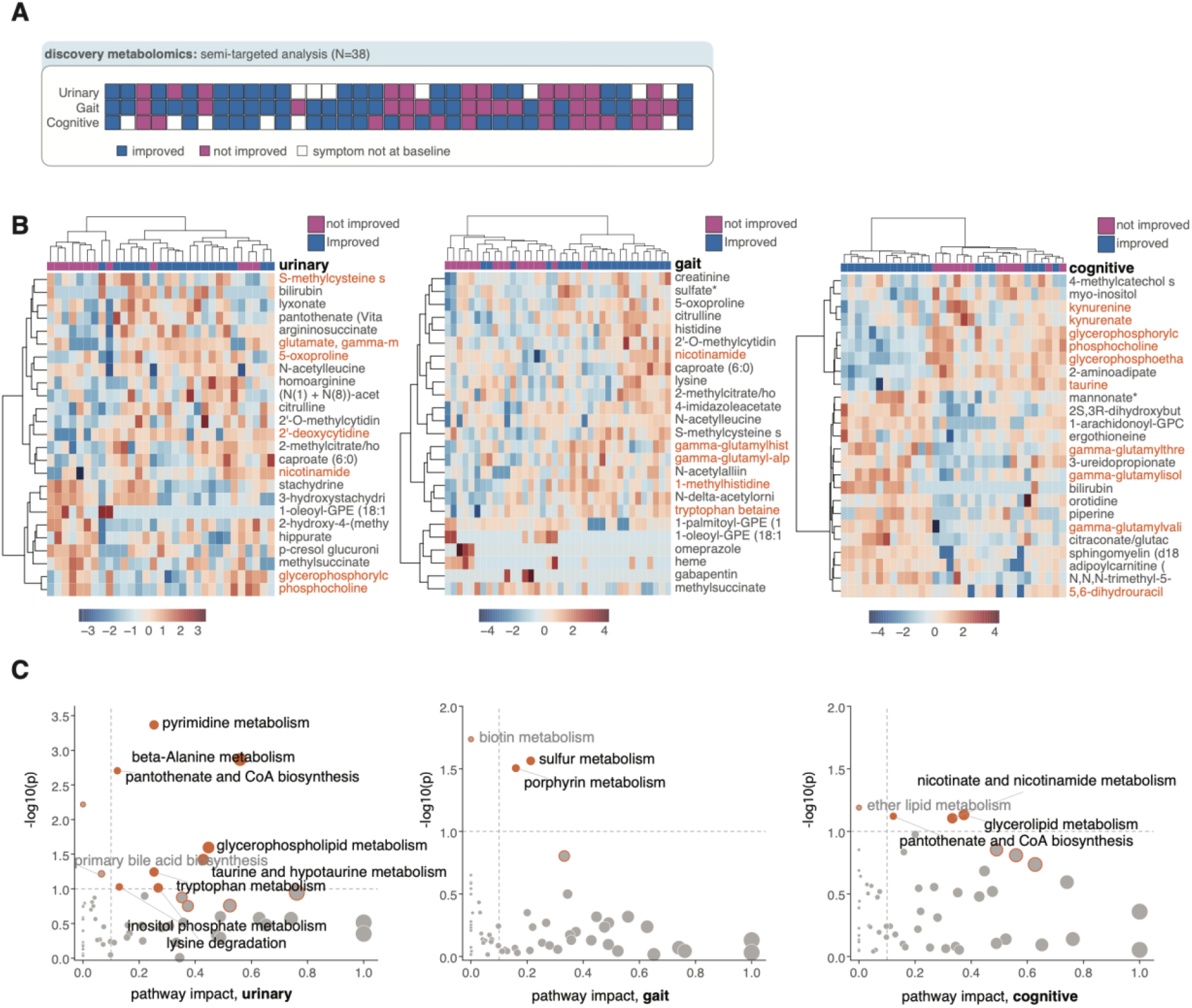
Discovery metabolomics of ventricular CSF. (**A**) Semi-targeted cohort: 42 patients profiled, of whom 38 had a 3-month outcome in at least one domain. Each column is one patient; rows show urinary, gait and cognitive outcome (improved, blue; not improved, magenta; symptom absent at baseline, white). Urinary: n = 31 (19 improved / 12 not improved); gait: n = 38 (22 / 16); cognition: n = 32 (20 / 12). 455 metabolite annotations were analyzed. (B) Heatmaps of the 25 metabolites with the largest group differences (t-test ranking) for each domain; rows, metabolites; columns, patients, annotated by outcome; colour, autoscaled log10 abundance; metabolites of interest in orange. (**C**) Pathway analysis (MetaboAnalyst, KEGG Homo sapiens; global test enrichment and relative-betweenness centrality topology, MetaboAnalyst defaults) for urinary, gait and cognitive outcomes. Each point is a pathway; x, pathway impact; y, −log10 raw p; point size, impact; orange, raw p < 0.1 (labelled); dashed lines, p = 0.1 and impact = 0.1. Pathways passing FDR < 0.05: urinary pyrimidine metabolism (FDR 0.026), β-alanine metabolism and pantothenate and CoA biosynthesis (both 0.039), which share the same measured intermediates; none for gait (lowest FDR 0.63) or cognition (0.82). *Alt text*: Three-part figure. A: grid of the 42 semi-targeted patients (columns) by urinary, gait and cognitive outcome (rows), each cell coloured blue for improved, magenta for not improved or white for symptom absent. B: three heatmaps, one per outcome, of 25 metabolites (rows) across patients (columns) grouped by outcome, coloured by relative abundance. C: three scatter plots of pathway analysis, one per outcome, plotting pathway impact against statistical significance; only urinary pyrimidine, beta-alanine and pantothenate/coenzyme A pathways pass the false discovery rate threshold.

To identify metabolites associated with clinical response to shunt surgery, we compared improved (defined as shunt responders) and non-improved (shunt non-responders) patients within each symptom domain (**Supplementary Fig. 2B; Fig. 2A**). No individual metabolite remained significant after multiple-hypothesis correction, and this discovery stage was used for hypothesis generation. Unsupervised clustering of the 25 metabolites with the largest group differences (t-test ranking) showed partial separation between improved and non-improved patients alongside substantial inter-individual variability (**Fig. 2B**).

To assess whether these trends reflected coordinated biochemical changes rather than isolated metabolite fluctuations, we performed pathway-level analysis (KEGG, *Homo sapiens*) comparing shunt responders and non-responders (**Fig. 2C**). For urinary outcomes, pyrimidine metabolism (FDR = 0.026), β-alanine metabolism and pantothenate and CoA biosynthesis (both FDR = 0.039) passed correction; these pathways share the same measured intermediates (uracil, dihydrouracil, 3-ureidopropionate, β-alanine, pantothenate) and therefore represent one route rather than three independent findings. No pathway passed correction for gait or cognitive outcomes (lowest FDR 0.63 and 0.82, respectively).

To evaluate these pathway-level signals at the level of individual metabolites, we examined the direction and size of metabolite differences in the corresponding pathways using the same covariate-adjusted models as in the targeted analysis^63^ (**Supplementary Fig. 2C–D**). In the pyrimidine-catabolism route, dihydrouracil (fold change improved/not improved 1.92, p < 0.001) and 3-ureidopropionate (1.47, p = 0.031) were higher in patients with cognitive improvement, and pantothenate was higher in patients with urinary improvement (1.39, p = 0.030), whereas uracil and β-alanine did not differ. Phosphocholine and glycerophosphocholine were lower in patients who improved in cognition (0.74, p = 0.003; 0.65, p = 0.014) and in urinary function (0.80, p = 0.010; 0.71, p = 0.030), as was taurine (cognition 0.76, p = 0.008; urinary 0.87, p = 0.018). In the tryptophan pathway, kynurenine and kynurenic acid were lower in patients with cognitive improvement (0.78, p = 0.014; 0.74, p = 0.019). No metabolite in these pathways differed for gait. These per-metabolite comparisons are nominal and exploratory; quinolinate and hypotaurine were detected in fewer than half of the samples and were not tested.

### Defining pathway indices from the discovery modules

Three modules recurred in the discovery analysis: one-carbon and sulfur-amino-acid metabolism together with its choline branch, the kynurenine branch of tryptophan metabolism, and alternative carbon sources (short-chain fatty acids, sugars and the pyrimidine–β-alanine–CoA route). To test these modules in the expanded targeted cohort without testing many correlated metabolites one at a time, we summarized each as a pathway index (Methods). The one-carbon/redox index (P1) combines the six metabolites of the first module measured on the targeted panel, cystathionine, 2-hydroxybutyrate, 2-aminobutyrate, 5-methyltetrahydrofolate, phosphocholine and phosphoethanolamine, with signs fixed from the position of each metabolite in the pathway; the kynurenine index (P2) combines tryptophan, kynurenine and their ratio; and eleven ratios of adjacent pathway metabolites, including a short-chain fatty acid/hexose ratio, served as secondary measures. The pyrimidine–β-alanine–CoA route could not be indexed because its intermediates did not survive quality control or were not detected on the targeted panel. The pathways were taken from the discovery cohort, and the choice of component metabolites was also informed by an initial exploratory analysis of the targeted data that had highlighted the same metabolites. Index definitions, covariates and multiple-testing families were then fixed in a statistical analysis plan, on a locked dataset, before the outcome associations reported below were computed. The index tests are therefore prespecified with respect to the analysis reported here, but not independent of the data that motivated them, and they require validation in an independent cohort.

### Targeted metabolomics identifies metabolic pathways associated with treatment responsiveness

We applied a targeted metabolomics assay focused on polar metabolites, leveraging an in-house library of approximately 500 compounds.^64^ The targeted cohort consisted of 55 ventricular CSF samples (31 from cohort 1 and 24 from cohort 2; **Fig. 3A**), in which 155 distinct metabolites passed quality control. No sample was removed on the basis of principal component analysis (**Supplementary Fig. 3A**).

**Figure 3.**
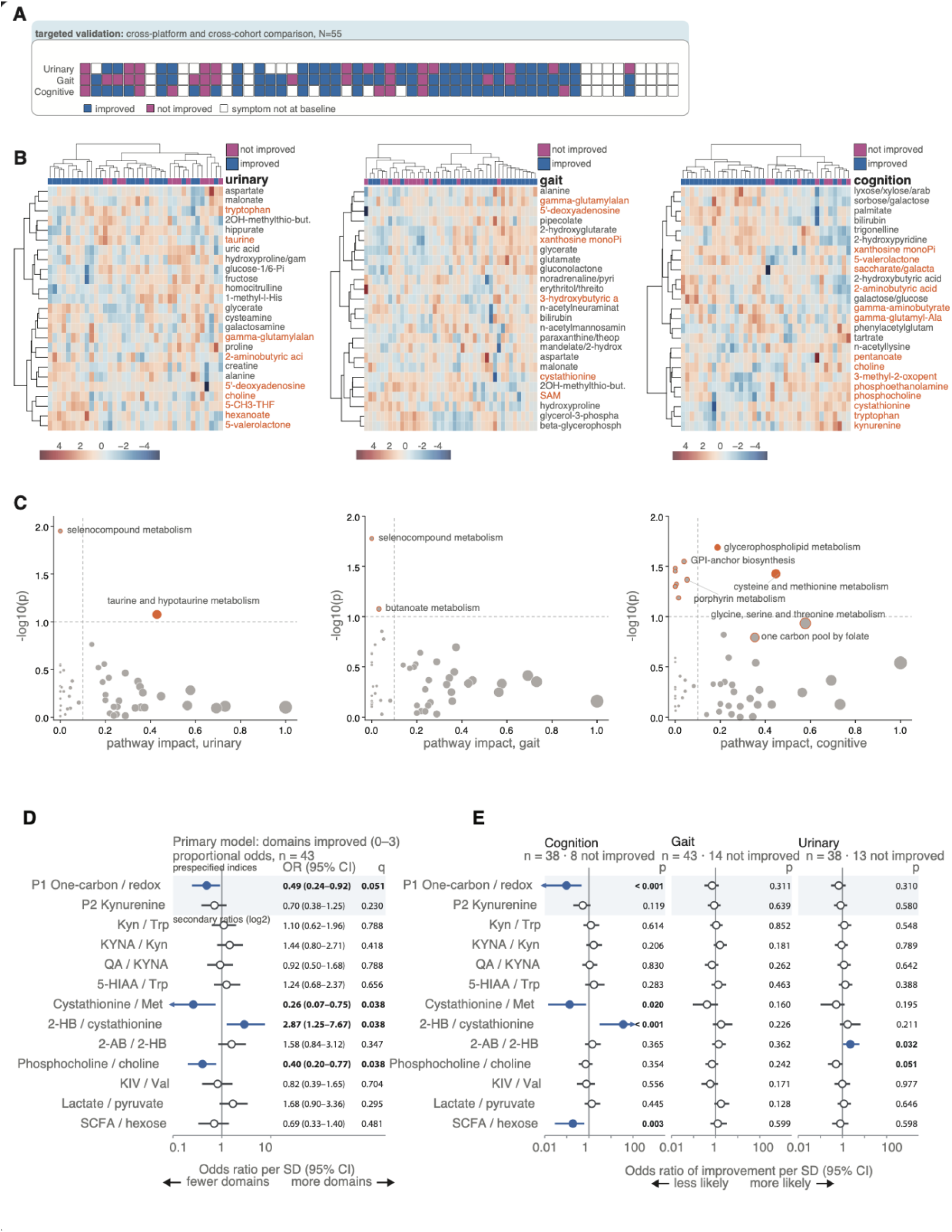
Targeted metabolomics of ventricular CSF and prespecified pathway indices. (**A**) Targeted cohort (n = 55 ventricular CSF samples passing quality control; 31 cohort 1, 24 cohort 2), coloured as in Fig. 2A. Urinary: n = 38 (25 / 13); gait: n = 43 (29 / 14); cognition: n = 38 (30 / 8). No sample was excluded on the basis of principal component analysis. (**B**) Heatmaps of the top 25 metabolites per domain, as in Fig. 2B (descriptive). (**C**) Pathway analysis as in Fig. 2C. No pathway passed FDR < 0.05 (lowest FDR 0.26, cognition); highest-ranked: cognition, glycerophospholipid metabolism (p = 0.020) and cysteine and methionine metabolism (p = 0.037, impact 0.45); urinary, taurine and hypotaurine metabolism (p = 0.084); gait, butanoate metabolism (p = 0.084); selenocompound metabolism (urinary, gait) rests on a single compound (impact 0). (D) Prespecified indices, primary model: proportional-odds regression of the number of domains improved (0–3) on each index (per SD), adjusted for age, sex and **enrollment** cohort (n = 43; 7 / 5 / 14 / 17 patients with 0 / 1 / 2 / 3 domains improved). Odds ratio per SD with profile-likelihood 95% CI, likelihood-ratio p and Benjamini–Hochberg FDR (q) within the primary family (P1 one-carbon/redox, P2 kynurenine; shaded) and the secondary family (eleven log2 ratios). Odds ratios below 1 indicate that higher index values accompany fewer domains improved. (E) Per-domain models (exploratory), adjusted as in (D): Firth-penalized logistic regression with profile penalized-likelihood 95% CI and p; cognition n = 38 (8 not improved), gait n = 43 (14 not improved), urinary n = 38 (13 not improved). In (**D–E**), filled blue symbols, p < 0.05; bold q, q < **0.10 in (D), p < 0.10 in (E);** arrowheads, interval extends beyond the axis. Primary family: no index reaches q < 0.05 (P1 q = 0.051); secondary family: cystathionine/methionine, 2-hydroxybutyrate/cystathionine and phosphocholine/choline, q = 0.038. *Alt text:* Five-part figure. A: grid of the 55 targeted-cohort patients by outcome, coloured as in Figure 2. B: heatmaps of the top 25 metabolites per outcome. C: pathway analysis scatter plots per outcome, with no pathway passing the false discovery rate threshold. D: forest plot of odds ratios with 95% confidence intervals for the two prespecified indices and eleven ratios against the number of domains improved; the one-carbon/redox index lies below 1. E: forest plots of the same indices for cognition, gait and urinary outcomes separately, with the one-carbon/redox index furthest below 1 for cognition.

We first assessed individual metabolite associations with clinical response. Fold-change volcano plots showed nominal differences across multiple metabolites, none significant after correction for multiple testing (**Supplementary Fig. 3B**). In covariate-adjusted models (age, sex and enrolment cohort; Methods), the number of nominally significant metabolites was close to that expected by chance (number of domains improved, 7 of 129 endogenous metabolites; cognition 9; gait 1; urinary 3; 6.5 expected), and no metabolite reached a false discovery rate below 0.05 (**Supplementary Fig. 3C**). The lowest was phosphocholine for cognition (odds ratio 0.11 per SD, FDR = 0.08). Unsupervised clustering of the top 25 metabolites ranked by group difference showed partial separation between shunt responders and non-responders (**Fig. 3B**).

To place these metabolite-level trends into a broader biochemical context, we performed pathway analysis (KEGG, *Homo sapiens*; **Fig. 3C**). For cognitive outcomes, glycerophospholipid metabolism (p = 0.020) and cysteine and methionine metabolism (p = 0.037; impact 0.45) ranked highest; for urinary outcomes, taurine and hypotaurine metabolism (p = 0.084); for gait, butanoate metabolism (p = 0.084). Selenocompound metabolism ranked first for urinary and gait outcomes but rests on a single compound (impact 0). No pathway passed correction for multiple testing (lowest FDR 0.26), so these results are descriptive. The cognitive signal centered on choline and sulfur-amino-acid metabolism, in line with the semi-targeted findings.

### Prespecified pathway indices identify a one-carbon/redox association with recovery

We next tested the prespecified indices (**Fig. 3D–E**). For the primary outcome, a higher one-carbon/redox index (P1) was associated with fewer domains improved (odds ratio 0.49 per SD, 95% CI 0.24–0.92, p = 0.026, FDR = 0.051 across the two primary indices; n = 43, distribution of 0/1/2/3 domains improved 7/5/14/17), whereas the kynurenine index (P2) was not (0.70, 0.38–1.25, p = 0.23). Among the eleven secondary ratios, those along the transsulfuration and choline branches pointed in the same direction (phosphocholine/choline 0.40 per SD, p = 0.006; cystathionine/methionine 0.26, p = 0.009; 2-hydroxybutyrate/cystathionine 2.87, p = 0.010; FDR 0.038 for each). By domain, the association was strongest for cognition (P1 odds ratio 0.10 per SD, 95% CI 0.004–0.46, p = 0.0006; Firth-penalized model, 8 non-improved patients) and absent for gait and urinary outcomes (p = 0.31 for both). The short-chain fatty acid/hexose ratio was also associated with cognitive improvement (0.19 per SD, p = 0.003), although not with the primary outcome (p = 0.31). Across sensitivity analyses the P1 odds ratio ranged from 0.43 to 0.56 (Supplementary Table 1b); it remained below p = 0.05 when patients who worsened were removed (0.50, p = 0.047), after removing three samples with flagged normalization factors (0.50, p = 0.034), after adjustment for body mass index (0.48, p = 0.025) and in a quasi-binomial model (0.46, p = 0.032), but not in iNPH patients only (0.56, p = 0.073), in patients with all three domains scored (0.50, p = 0.061) or in cohort 2 alone (0.43, n = 16, p = 0.17); no P1 value had |robust z| > 3.5. Individually, the metabolites that ranked highest for cognition in the adjusted scan were the P1 components themselves, with directions matching the signs fixed in the analysis plan.

Finally, we evaluated metabolites previously associated with NPH disease status or shunt responsiveness^65–68^ with the same adjusted models on both platforms (**Table 2**). Only targeted 2-hydroxybutyrate was nominally associated with an outcome (cognition, higher in improved patients, odds ratio 3.11 per SD, p = 0.025, FDR = 0.55). Serine and 2-hydroxybutyrate associations with cognition reported in an exploratory, unadjusted analysis of the semi-targeted data were not reproduced after covariate adjustment. These results indicate limited overlap between previously reported NPH metabolites and outcome-associated signals in this cohort.

**Table 2.** Previously reported NPH-associated metabolites tested in the present cohort. Each metabolite was tested on each platform where measured with the covariate-adjusted models used for the per-metabolite analysis (number of domains improved, and each domain). Table body to be replaced with the compact sheet of Table2_literature_metabolites_v1.2.xlsx. Abbreviations: HVA, homovanillic acid; 5-HIAA, 5-hydroxyindoleacetic acid; NS, not significant (all p ≥ 0.05); NPH, normal pressure hydrocephalus. * Directionality reported in the cited literature (↑ increased, ↓ decreased in disease or among patients demonstrating postoperative improvement, as specified)

| **Metabolite** | **Reported context** | **Direction in literature\*** | **Semi-targeted cohort, *P* value** | **Targeted cohort, *P* value** | **PMID** |
|---|---|---|---|---|---|
| Homovanillic acid | NPH disease | ↑ | NA | NA | <sup>66</sup> |
| HVA/5-HIAA ratio | NPH disease | ↑ | NA | NA | <sup>66</sup> |
| Serine | NPH disease | ↑ | 0.0410 | NS | <sup>67</sup> |
| Glutamic acid | NPH disease | ↑ | NA | NS | <sup>67</sup> |
| 2-Hydroxybutyrate | NPH disease | ↑ | 0.0396 | 0.0482† | <sup>65</sup> |
| Glycerate | NPH disease | ↓ | NA | 0.0270‡ | <sup>65</sup> |
| N-Acetylneuraminate | NPH disease | ↓ | NA | NS | <sup>65</sup> |
| S-Adenosylmethionine | Shunt improvement | ↓ | NA | 0.0490§ | <sup>68</sup> |
| Succinate | Shunt improvement | ↑ | NA | 0.0340§ | <sup>68</sup> |
| Citrate | Shunt improvement | ↑ | NA | NS | <sup>68</sup> |

### Multivariate prediction points to a modest cognitive signal

We next asked how well the metabolite profile predicts outcome in individual patients. In small cohorts, performance estimated on the same data used to select and tune a model is optimistic,^51^ so all prediction analyses used nested cross-validation, with all preprocessing, feature selection and tuning fitted inside each training fold, and significance was assessed against a permutation null of the entire pipeline (1,000 label permutations) (Methods). We used two strategies (Fig. 4A; Supplementary **Fig.** 4A): the leave-one-out pipeline of our initial analysis, re-run in nested form, and the locked protocol (outer 5-fold, repeated 20 times; inner 5-fold), in which ridge-penalized logistic regression on all metabolites was prespecified as the primary model and a linear support vector machine (SVM) and an RBF-kernel SVM, each using the top-ranked metabolites by logistic-regression weight, were comparators.

**Figure 4.**
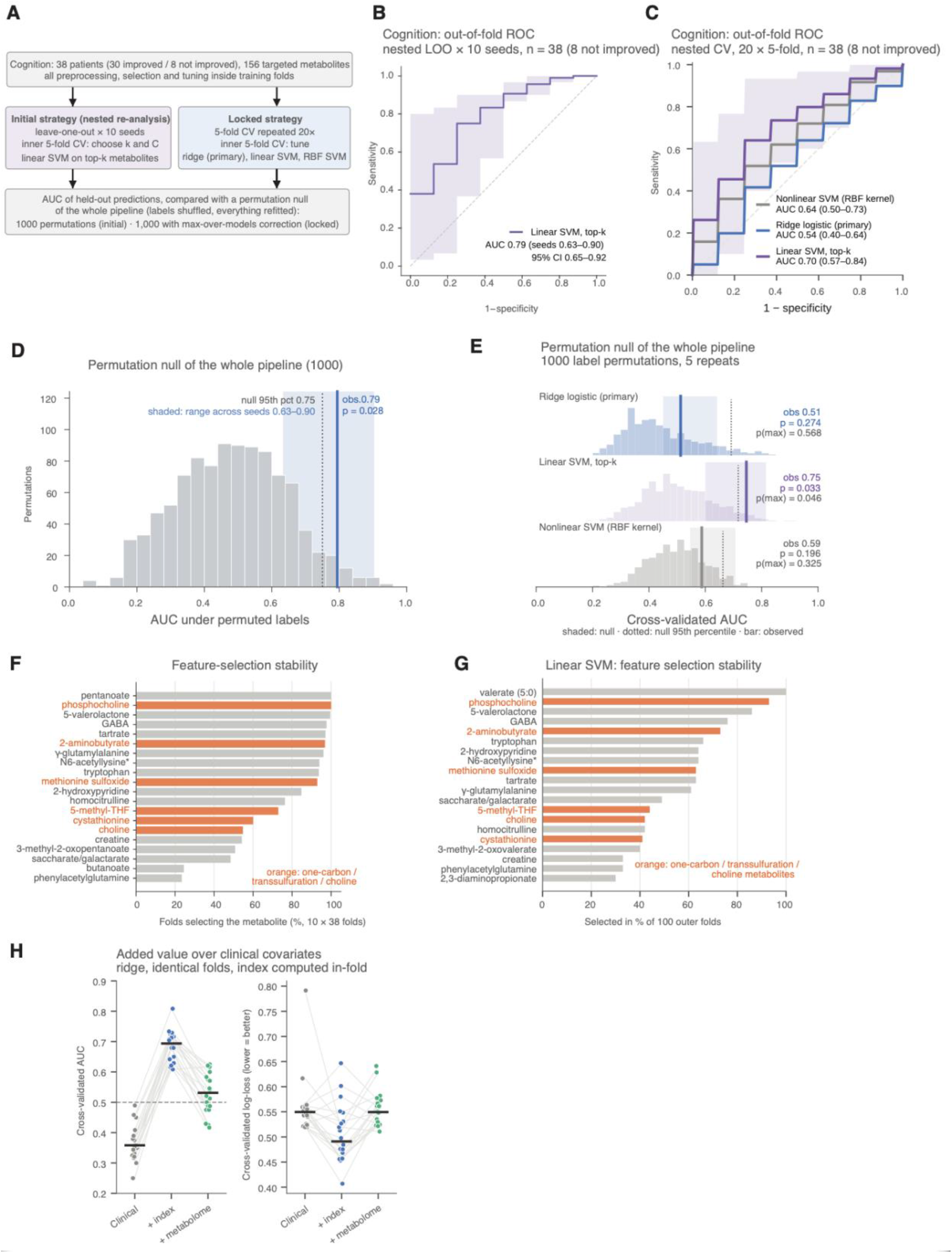
Prediction of cognitive improvement under two nested cross-validation strategies. Cognition, n = 38 (30 improved, 8 not improved), 156 targeted metabolite columns (155 distinct metabolites); all preprocessing, feature selection and tuning fitted within training folds. (A) The two strategies (details in Supplementary Fig. 4A). **(B, D, F)** Leave-one-out pipeline of the initial analysis, re-run in nested form after the analysis lock. **(C, E, G, H)** Locked protocol: outer 5-fold cross-validation repeated 20 times with inner 5-fold tuning. **(B)** Out-of-fold ROC curve of the linear SVM on the top-k metabolites; mean curve across 10 seeds (seeds change only the inner split), shaded band, range across seeds; dashed diagonal, chance. AUC 0.79 (range across seeds 0.63–0.90; 95% patient-bootstrap CI 0.65–0.92). **(C)** Out-of-fold ROC curves for ridge logistic regression (prespecified primary model), linear SVM on the top-k metabolites, and RBF-kernel (nonlinear) SVM on the top-k metabolites; mean curve per model across repeats, shaded band, range across repeats for the linear SVM; dashed diagonal, chance. Median AUC (range across repeats): ridge 0.54 (0.40–0.64), linear SVM 0.70 (0.57–0.84), RBF-kernel SVM 0.64 (0.50–0.73). **(D)** Permutation null of the whole leave-one-out pipeline: AUC distribution from 1,000 label permutations (grey), null 95th percentile 0.75 **(dotted),** observed mean AUC **(line) and range of the observed AUC across seeds (shaded box);** p = 0.028 for this pipeline alone. **(E)** Permutation null of the whole locked pipeline: AUC distribution from 1,000 label permutations (5 outer repeats each; shaded) with the null 95th percentile (dotted) and the observed AUC at the same 5 repeats **(bar); box, range of the observed AUC across these 5 repeats.** Observed AUC, permutation p and p after max-over-models correction (p(max)): ridge 0.51, 0.274, 0.568; linear SVM 0.75, 0.033, 0.046; RBF-kernel SVM 0.59, 0.196, 0.325. **(F, G)** Feature-selection **stability:** percentage of outer folds in which each metabolite was selected (top 20), for the leave-one-out pipeline (F; 380 folds, 10 seeds × 38) and the linear SVM of the locked protocol **(G;** 100 folds); orange, one-carbon, transsulfuration and choline metabolites. **(H)** Incremental value over clinical covariates (ridge, identical folds): clinical covariates (age, sex, cohort), plus the prespecified indices P1 and P2 recomputed within each training fold (+ index), and plus the full metabolome (+ metabolome). Left, cross-validated AUC; right, cross-validated log-loss (lower is better). Points, the 20 repeats (lines connect the same repeat); bars, medians. Median log-loss 0.550, 0.491 and 0.549; the clinical-only model shrinks to near-constant predictions, so its AUC falls below 0.5 (0.36) and log-loss is the fair comparison. The higher leave-one-out estimate reflects the cross-validation scheme, under which the permutation null also rises (0.75 vs 0.72); the two estimates are consistent. *Alt text*: Eight-part figure on predicting cognitive improvement with two cross-validation strategies. A, diagram of the two strategies. B and C, ROC curves: the leave-one-out strategy (B; linear support vector machine, AUC 0.79) and the locked strategy (C; three models, linear support vector machine highest, median AUC 0.70), all above the chance diagonal. D and E, histograms of the AUC under permuted labels with the observed values marked, above the 95th percentile for both linear support vector machines. F and G, bar charts of how often the 20 most frequently selected metabolites were chosen in each strategy, with one-carbon, transsulfuration and choline metabolites highlighted. H, paired dot plots comparing clinical, clinical plus index and clinical plus metabolome models by AUC and log-loss, where adding the indices lowers the log-loss.

For cognition, the leave-one-out pipeline of our initial analysis, re-run in nested form, distinguished improved from non-improved patients with a mean AUC of 0.79 across 10 seeds (range 0.63–0.90; 95% CI 0.65–0.92) (Fig. 4B), above its permutation null (95th percentile 0.75; p = 0.028) (Fig. 4D). The metabolites selected most often included phosphocholine, 2-aminobutyrate and methionine sulfoxide (each in more than 90% of folds), together with 5-methyltetrahydrofolate, cystathionine and choline, and pentanoate, 5-valerolactone, γ-aminobutyrate, tartrate and tryptophan (Fig. 4F).

In the locked protocol, the sparse linear SVM distinguished improved from non-improved patients with a median cross-validated AUC of 0.70 (range across repeats 0.57–0.84), above the permutation null after accounting for the choice among three models (p = 0.046; p = 0.033 for this model alone) (**Fig.** 4C, E). Among the metabolites it selected most often were one-carbon, transsulfuration and choline metabolites, phosphocholine, 2-aminobutyrate, methionine sulfoxide, 5-methyltetrahydrofolate, choline and cystathionine, together with pentanoate, 5-valerolactone and γ-aminobutyrate (**Fig.** 4G). The prespecified ridge model, which includes all metabolites, achieved an AUC of 0.54 (0.40–0.64; p = 0.27), while the RBF-kernel SVM reached 0.64 (0.50–0.73; p = 0.20). At this sample size, none of the models provided reliable individual risk estimates. The ridge model produced nearly identical predicted risks across patients, close to the overall rate of non-improvement in the cohort (calibration slope −0.07; Brier score 0.174 vs 0.166 for predicting the prevalence).^69^ Similarly, at the thresholds defined in advance, the linear SVM identified only a small proportion of non-improvers (sensitivity 0.13, specificity 0.93) (**Supplementary Fig. 5A–C**). The cognitive signal is therefore detectable across patients but not yet precise enough to predict an individual’s outcome.

The two strategies give consistent estimates. The leave-one-out estimate is higher, but its permutation null is higher as well (95th percentile 0.75 vs 0.72 for the linear SVM in the locked protocol), so the difference reflects the cross-validation scheme rather than additional signal. Both are lower than the estimates of our initial exploratory analysis^70,71^ of the same data, in which the leave-one-out pipeline reached AUCs of 0.85 for cognition, 0.88 for gait and 0.73 for urinary outcomes (**Supplementary Table** 3d). In that pipeline, preprocessing and feature ranking were performed within each leave-one-out fold, but the number of metabolites was selected as the value giving the highest leave-one-out AUC, an approach that can lead to overly optimistic estimates in small samples^51^; applied to permuted outcome labels, the same procedure yielded AUCs of up to 0.84 (95th percentile of 100 permutations; **Supplementary Table** 3d). We therefore report the nested estimates, which are expected to be closer to the performance in new patients.

For gait and urinary outcomes, none of the models predicted clinical improvement significantly better than chance (gait median AUC 0.53–0.57; urinary 0.38–0.56; all permutation p ≥ 0.13) (**Supplementary Fig.** 7; **Supplementary Table 3**). The leave-one-out pipeline gave mean AUCs of 0.65 for gait (range across seeds 0.57–0.78; null 95th percentile 0.71; p = 0.12) and 0.63 for urinary outcome (0.56–0.72; null 95th percentile 0.73; p = 0.16) (Supplementary Fig. 6; Supplementary Table 3d). We then asked whether metabolite information adds to clinical covariates, using the same folds for all models (**Fig.** 4H). For cognition, adding the two prespecified indices, recomputed within each training fold, to age, sex and enrolment cohort reduced the cross-validated log-loss from 0.550 to 0.491, whereas adding the full metabolome did not (0.549). Because the clinical-only model was shrunk to near-constant predictions, its cross-validated AUC fell below 0.5 (0.36) and AUC differences overstate the gain; log-loss was therefore used for this comparison. In an exploratory analysis added after the selection frequencies had been inspected, further adding the short-chain fatty acid/hexose ratio gave a log-loss of 0.486 (**Supplementary Table 3**). For gait and urinary outcomes, neither the indices nor the metabolome improved on clinical covariates. For cognition, the prespecified indices therefore carried the predictive signal more efficiently than the full metabolome.

### Metabolite and transcript signals converge on one-carbon, choline and alternative carbon metabolism

We summarized the metabolite-level results on pathway maps, using one fold change (improved/not improved) and one adjusted test throughout (**Fig. 5A–C**). The metabolites that carried the prespecified index result, the adjusted per-metabolite scan and the most frequent features of the linear SVM overlapped: cystathionine, phosphocholine, phosphoethanolamine, 2-aminobutyrate, 2-hydroxybutyrate and pentanoate recurred across these analyses for cognition.

**Figure 5.**
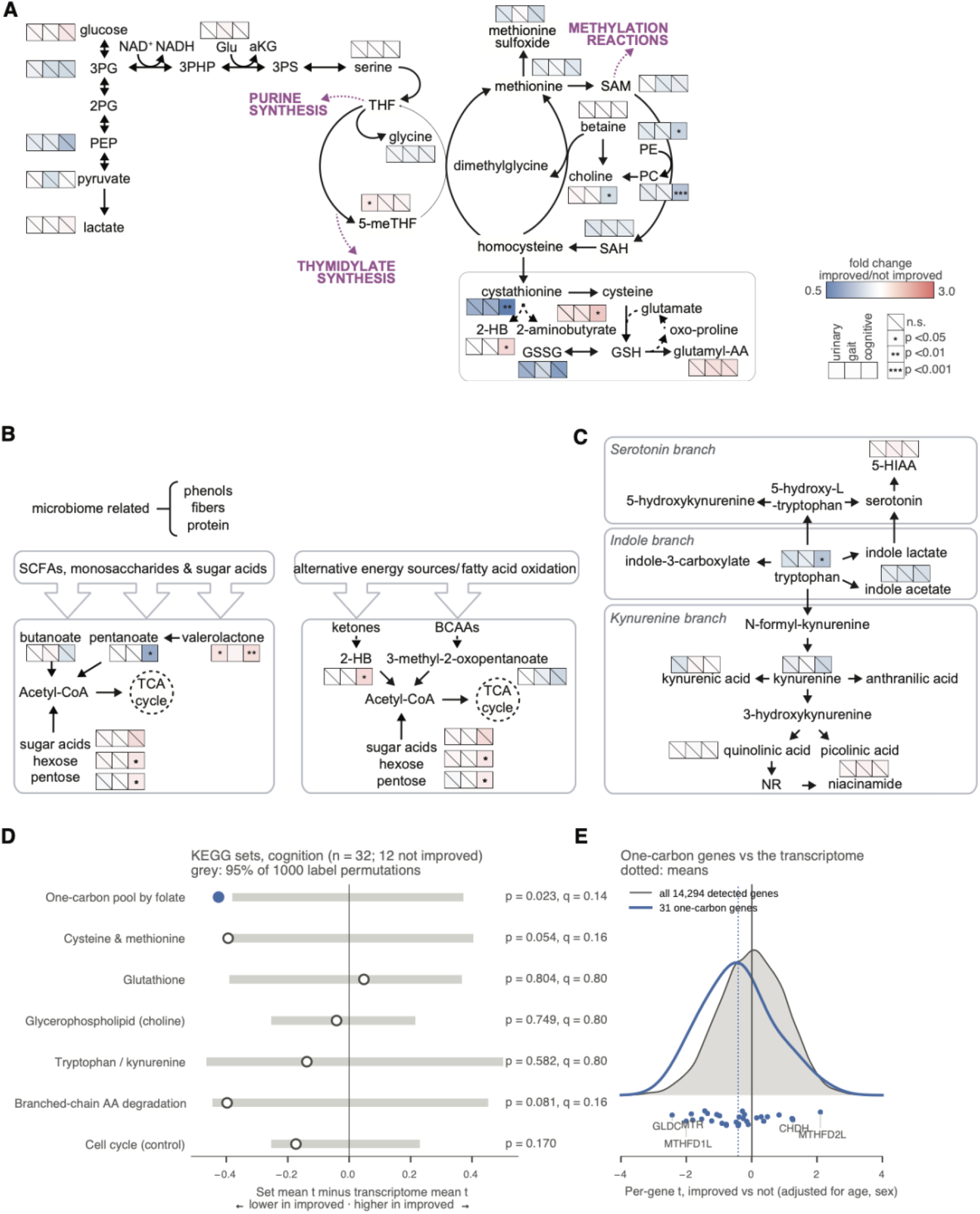
Metabolite and transcript differences between improved and non-improved patients in one-carbon, alternative carbon and tryptophan metabolism. (**A**) One-carbon metabolism, the methionine cycle, transsulfuration and glutathione synthesis (targeted ventricular CSF). For each metabolite, three squares show urinary, gait and cognitive outcomes; colour, fold change improved/not improved (blue, lower in improved; red, higher in improved; scale [0.5–3.0]); stars, p from the covariate-adjusted per-metabolite models (*p < 0.05, **p < 0.01, ***p < 0.001); slanted line, not significant. Abbreviations: 2-HB, 2-hydroxybutyrate; 2PG, 2-phosphoglycerate; 3PG, 3-phosphoglycerate; 3PHP, 3-phosphohydroxypyruvate; 3PS, 3-phosphoserine; 5-meTHF, 5-methyltetrahydrofolate; aKG, α-ketoglutarate; Glu, glutamate; glutamyl-AA, γ-glutamyl amino acid; GSH, reduced glutathione; GSSG, oxidized glutathione; PC, phosphocholine; PE, phosphoethanolamine; PEP, phosphoenolpyruvate; SAH, S-adenosylhomocysteine; SAM, S-adenosylmethionine; THF, tetrahydrofolate. (**B**) As in (A), for microbiome-related and alternative energy-source pathways (short-chain fatty acids, monosaccharides and sugar acids; ketones, branched-chain amino acids and fatty acid oxidation). SCFA, short-chain fatty acids; BCAAs, branched-chain amino acids. (**C**) As in (A), for the serotonin, indole and kynurenine branches of tryptophan metabolism. 5-HIAA, 5-hydroxyindoleacetic acid; NR, nicotinamide riboside. Nodes without squares were not measured on the targeted platform. (**D**) Competitive gene-set test in CSF extracellular-vesicle RNA (cognition, n = 32, 12 not improved; cohort 1): for each KEGG set, mean outcome t-statistic of the set minus that of all 14,294 detected genes (per-gene linear model adjusted for age and sex); grey bars, central 95% of 1,000 label permutations; filled, p < 0.05; p and FDR across the six metabolic sets; cell cycle, non-metabolic control. Negative values, lower expression in improved patients. (E) Per-gene t-statistics of the 31 one-carbon pool genes (blue) against all detected genes (grey density); dotted lines, means; selected genes labelled. *Alt text*: Five-part figure. A to C: pathway diagrams of one-carbon and transsulfuration metabolism, alternative carbon sources, and tryptophan metabolism; beside each measured metabolite, three small squares show the urinary, gait and cognitive fold change, blue when lower and red when higher in improved patients, with stars marking significance. D: bar chart of gene-set statistics for six metabolic KEGG pathways and a cell-cycle control, with permutation intervals. E: density plot of per-gene statistics for all detected genes with the one-carbon pool genes overlaid.

In one-carbon and transsulfuration metabolism (**Fig. 5A**), patients with cognitive improvement had lower cystathionine (fold change 0.47, p = 0.008), phosphocholine (0.77, p = 0.001) and phosphoethanolamine (0.82, p = 0.029), and higher 2-aminobutyrate (1.69, p = 0.016) and 2-hydroxybutyrate (1.46, p = 0.025). Urinary improvement was accompanied by higher 5-methyltetrahydrofolate (1.27, p = 0.031). In alternative carbon metabolism (**Fig. 5B**), pentanoate was lower (0.67, p = 0.016) and 5-valerolactone (1.39, p = 0.010) and pentoses (1.23, p = 0.034) were higher in patients with cognitive improvement. In the tryptophan pathway (**Fig. 5C**), tryptophan was lower in patients with cognitive improvement (0.79, p = 0.025), while kynurenine and its downstream metabolites did not differ on the targeted platform. Semi-targeted data showed the same directions for phosphocholine, glycerophosphocholine and the kynurenine branch (**Supplementary Fig. 2C–D**). These per-metabolite comparisons are nominal and serve to describe the index result, not as independent tests.

Next, we asked whether metabolite-associated signals were consistent with transcriptional changes in CSF extracellular vesicles from the same cohort 1 patients.^43^ We first tested 19 enzyme–metabolite pairs with expected directions, fixed before the transcriptomic matrix was opened; seven could not be tested because the genes were detected in fewer than half of the samples, and none of the remaining twelve was significant (all FDR ≥ 0.54; **Supplementary Table 4**). We then asked a pathway-level question: whether outcome-associated transcripts are concentrated in one-carbon metabolism, using KEGG gene sets fixed before analysis and a competitive test against all 14,294 detected genes with patient-label permutation^54^ (Methods). For cognition (n = 32), transcripts of the one-carbon pool by folate set were shifted towards higher expression in non-improved patients (permutation p = 0.023; FDR = 0.14 across six metabolic sets), with cysteine and methionine metabolism (p = 0.054) and branched-chain amino acid degradation (p = 0.081) in the same direction, whereas glutathione, glycerophospholipid and tryptophan sets and a cell-cycle control set were not (**Fig. 5D–E**). The shift was distributed across the set rather than driven by single genes (2 of 31 genes individually nominal), it was not significant after correction, and it was not seen for the number of domains improved (p = 0.16). Its direction matches the metabolite result: a higher one-carbon/redox index and higher one-carbon transcript levels both accompany less cognitive improvement. Because these are the same patients as the semi-targeted discovery cohort, this is consistent across molecular layers rather than independent replication.

Together, these results point to coordinated differences in one-carbon, choline and alternative carbon metabolism in intraoperative CSF of patients who recover cognitive function, captured by the prespecified one-carbon/redox index and mirrored, in the same direction, by one-carbon transcripts in CSF extracellular vesicles.

### Recovery-associated indices are only partly reflected in lumbar CSF and plasma

To assess whether the molecular signature obtained from intraoperative ventricular samples is also reflected in more clinically accessible lumbar CSF or plasma, we analyzed the targeted dataset across biofluids (**Fig. 6A** and **Supplementary Fig. 1C**): 55 ventricular CSF samples (31 in cohort 1 and 24 in cohort 2), 29 lumbar CSF samples and 33 plasma samples. Twelve patients had matched ventricular CSF, lumbar CSF and plasma, and 24 had matched ventricular CSF and plasma. Lumbar CSF was collected at the diagnostic tap test and ventricular CSF at surgery, on different days.

**Figure 6.**
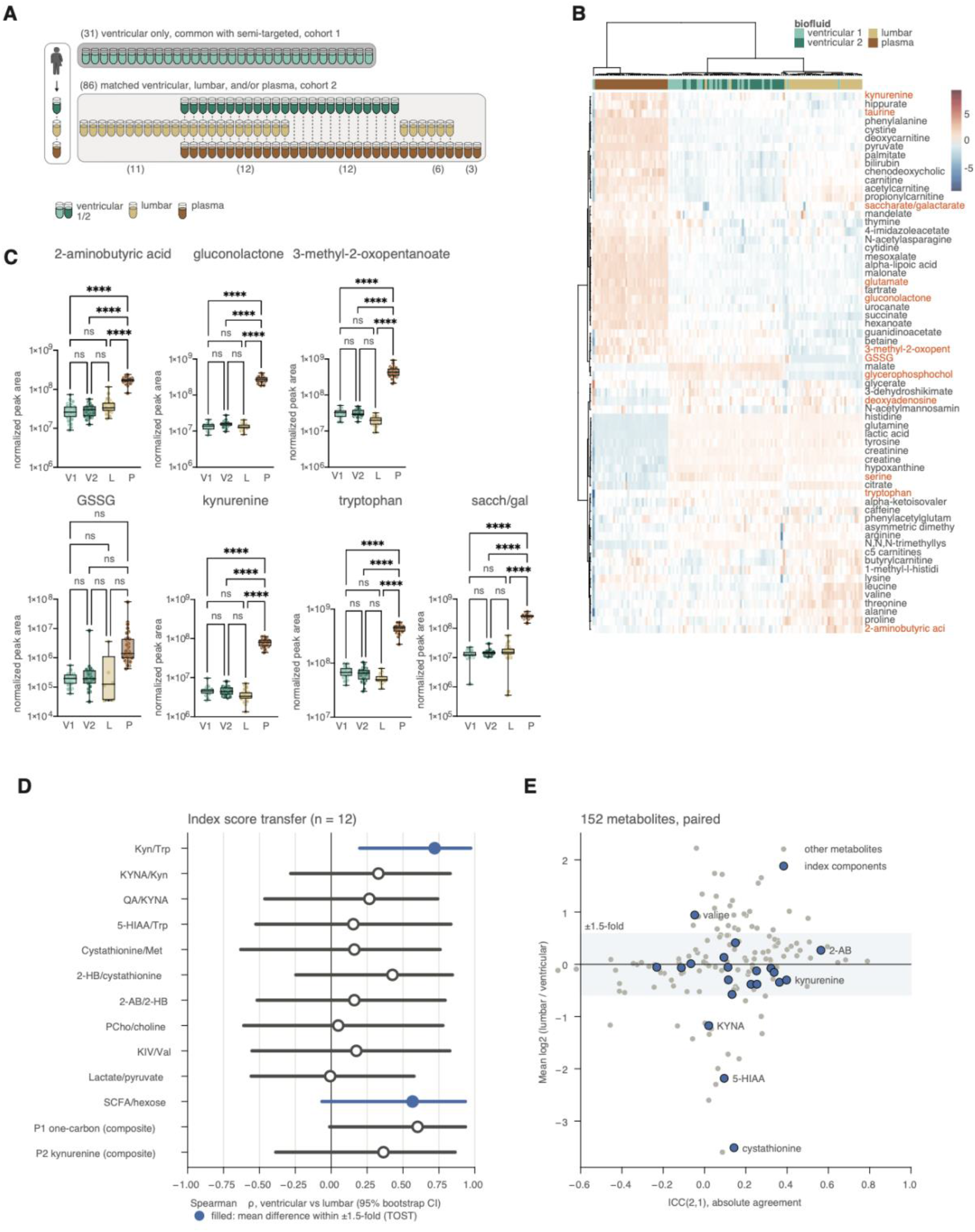
Comparative analysis of targeted metabolite profiles across ventricular, lumbar, and plasma samples. **(A)** Samples analyzed on the targeted platform, regardless of clinical follow-up: ventricular CSF cohort 1 (n = 31, also measured on the semi-targeted platform), ventricular CSF cohort 2 (n = 24), lumbar CSF (n = 29) and plasma (n = 33). Matched sets: ventricular cohort 2, lumbar CSF and plasma, n = 12; ventricular cohort 2 and plasma, n = 12; lumbar CSF and plasma, n = 6; lumbar CSF only, n = 11; plasma only, n = 3. (**B**) Heatmap of targeted metabolites across ventricular cohorts 1 and 2, lumbar CSF and plasma (autoscaled log10; plasma values multiplied by 8.8 for the lower injected volume-equivalent); metabolites of interest in orange. (**C**) Normalized peak areas of selected metabolites by fluid (V1, V2, L, P); boxes, median and interquartile range; whiskers min-max; points, samples. One-way ANOVA with Šídák’s multiple comparisons test (ns, not significant; ****p < 0.0001). (**D**) Transfer of index values from ventricular to lumbar CSF in the 12 matched patients (outcome-blind): Spearman ρ with 95% bootstrap CI; filled, mean lumbar–ventricular difference within a prespecified ±1.5-fold margin (two one-sided tests); lumbar indices computed with the ventricular means, SDs and half-minimum values. (**E**) Per-metabolite agreement for 152 paired metabolites: intraclass correlation (ICC(2,1), absolute agreement) against mean log2(lumbar/ventricular); shaded band, ±1.5-fold; blue, index components. *Alt text*: Five-part figure comparing biofluids. A: diagram of the overlap between ventricular CSF from two cohorts, lumbar CSF and plasma samples. B: heatmap of targeted metabolites across the four sample groups, with plasma clearly distinct from CSF. C: box plots of selected metabolites by fluid with significance stars. D: dot plot of correlations with confidence intervals for the transfer of each index from ventricular to lumbar CSF in 12 matched patients. E: scatter plot of per-metabolite agreement between lumbar and ventricular CSF against their mean ratio, with index components highlighted.

Unsupervised clustering showed that metabolite profiles were highly consistent between the two ventricular CSF cohorts, which clustered together (**Fig. 6B**). To enable direct comparison across biofluids while accounting for matrix-dependent differences in signal intensity, plasma samples were injected at a lower plasma-equivalent volume to compensate for their higher metabolite concentrations, and plasma values were multiplied by 8.8 for all cross-fluid comparisons.^72,73^ Plasma remained clearly separated from both CSF types, whereas lumbar CSF clustered closer to ventricular CSF. After accounting for injected volume-equivalents, plasma concentrations were as expected higher across metabolites, with GSSG (oxidized glutathione) as a notable exception (**Fig. 6C**).

We then asked, without reference to outcome, whether the index values themselves transfer from ventricular to lumbar CSF in the 12 matched patients (**Fig. 6D–E**), using correlation and equivalence within a prespecified ±1.5-fold margin. Only the kynurenine/tryptophan ratio met both criteria (Spearman ρ = 0.72, 95% CI 0.20–0.97; equivalent within the margin). The one-carbon/redox index (ρ = 0.60, −0.01 to 0.94) and the short-chain fatty acid/hexose ratio (ρ = 0.57, −0.06 to 0.93; equivalent in level) correlated moderately with wide intervals, and most transsulfuration and choline ratios differed systematically between compartments. Recovery-associated metabolites showed partly concordant directions in lumbar CSF and weaker concordance in plasma (**Supplementary Fig.** 8), but with 20 lumbar and 24 plasma samples with outcomes these comparisons are descriptive.

Together, these results show that ventricular and lumbar CSF share overall metabolite profiles, but that among the prespecified indices only the kynurenine/tryptophan ratio transfers closely between them; whether the one-carbon/redox index can be measured in lumbar CSF requires a larger matched set.

## Discussion

In this study, we combined CSF metabolomics with a prespecified analysis on a locked dataset to investigate molecular states associated with neurological recovery after shunt surgery in NPH. A one-carbon/redox index, combining transsulfuration and choline metabolites, was associated with the number of symptom domains improved, most clearly for cognition, and transcripts of the one-carbon pathway in CSF extracellular vesicles from the same patients shifted in the same direction. Under nested cross-validation, metabolite-based prediction carried a modest signal for cognition, largely captured by the prespecified indices, but did not provide reliable predictions at the individual patient level; gait and urinary outcomes showed no robust metabolic signal.

To interpret these findings, we examined the biological pathways represented by the metabolite signals. Rather than isolated metabolite changes, the cognitive association was carried by several metabolites of one network, each with a modest effect, which is the situation a prespecified composite index is designed to detect and a metabolite-by-metabolite scan is not.

Patients with cognitive improvement had lower cystathionine, phosphocholine and phosphoethanolamine and higher 2-aminobutyrate and 2-hydroxybutyrate. Cystathionine is the transsulfuration intermediate between homocysteine and cysteine, 2-hydroxybutyrate and 2-aminobutyrate are by-products of cystathionine cleavage that rise with glutathione demand, and phosphocholine and phosphoethanolamine connect one-carbon metabolism to membrane phospholipid synthesis through methylation. The pattern is therefore consistent with differences in sulfur-amino-acid flux and in choline/phospholipid turnover prior to intervention, which are linked to redox capacity. Redox homeostasis is a key determinant of neuronal integrity and response to oxidative stress, and these findings are consistent with earlier observations in NPH, including lipid peroxidation and thiol imbalance,^74^ as well as broader evidence linking glutathione-related pathways to neurodegeneration and aging.^75–77^ The shift of one-carbon transcripts in the same direction supports a biological rather than an analytical origin, although it was not significant after correction.

The kynurenine pathway, prespecified as the second index, was not associated with the primary outcome, and differences in individual tryptophan-pathway metabolites were nominal. A recent targeted CSF study reported lower tryptophan in iNPH than in healthy controls and in other neurodegenerative diagnoses;^46^ how tryptophan levels relate to disease state on the one hand and to recovery on the other requires direct testing. Kynurenine metabolism is closely tied to neuroimmune signaling and microglial function,^78^ and prior CSF studies in NPH indicate inflammatory activation associated with disease state;^79,80^ whether kynurenine pathway activity relates to recovery remains open in this cohort.

Pathways related to alternative carbon utilization were also associated with cognitive outcome: the short-chain fatty acid/hexose ratio was associated with cognitive improvement, and pentanoate, a microbiome-derived short-chain fatty acid linked to immunomodulatory and epigenetic regulation,^81,82^ was lower and 5-valerolactone higher in patients who improved. These observations are consistent with emerging evidence connecting systemic metabolism, microbiome-derived metabolites, and neurological function.^83,84^ On the semi-targeted platform, the pyrimidine catabolism route to β-alanine and coenzyme A was the only pathway signal to pass correction (urinary outcome), and dihydrouracil was the strongest single semi-targeted metabolite for cognition; this route intersects one-carbon metabolism at thymidylate synthesis and warrants testing as a prespecified hypothesis.

Our analysis proceeded in two stages. In an initial exploratory stage, metabolites, pathways and classifiers were selected and evaluated on the same patients, as is common in discovery metabolomics; in a second stage, we fixed the dataset, the outcome coding, the indices and the analysis plan before testing. The second stage yielded lower but more robust performance estimates, consistent with the optimistic bias that arises when model settings are selected on the same cross-validation folds used to estimate performance,^51^ whereas the pathway-level association with cognition persisted under the stricter procedure, and two nested implementations, leave-one-out and repeated 5-fold, gave consistent estimates of a modest cognitive signal. With 8 non-improved patients for cognition, cohorts of this size can detect a signal but cannot establish or exclude a clinically useful predictor.^85^

Previous efforts to predict shunt outcome in idiopathic NPH have relied primarily on clinical testing and imaging-derived markers, with molecular readouts only recently incorporated into outcome analyses. Meta-analyses report substantial heterogeneity in outcome definitions and study design, limiting comparability and mechanistic interpretation.^86,87^ Biomarker studies have largely focused on diagnostic differentiation,^88^ and CSF amyloid-β and tau species distinguish NPH from Alzheimer’s disease but show limited specificity for predicting shunt responsiveness^89,90^; most recently, CSF p-tau181 was not associated with shunt response in 230 shunted patients across two assay platforms.^40^ More recent proteomic analyses demonstrate that baseline CSF protein states, including inflammatory and synaptic markers, associate with shunt responsiveness,^42,91–93^ supporting the concept that the preoperative molecular state reflects recovery-relevant biology, although a recent systematic review concluded that such proteins remain exploratory candidates pending validation in standardized prospective cohorts.^94^ Our previous transcriptomic profiling further linked shunt responsiveness to gene programs involved in mitochondrial function and proteostasis.^43^ In this context, the present study adds a complementary layer by capturing metabolite profiles that reflect an integrated functional output of these processes.

Recent work has highlighted persistent challenges in metabolomics and ML-based biomarker discovery, including variability in sample preparation, platform-dependent effects, and the risk of overfitting in high-dimensional datasets,^95,96^ and community recommendations now frame biomarker development as a staged process,^97,98^ from discovery and analytical validation to clinical validation of a locked model in an independent cohort, prospective evaluation and assessment of clinical utility.^99,100^ Measured against this framework, the present study spans discovery and early verification. Because the indices were informed by data from the same center and partly the same patients, and no external or prospective validation has yet been performed, the results should be read as evidence of a detectable, biologically coherent signal rather than as estimates of clinical accuracy.^96,101^

Beyond its relevance to NPH, this work illustrates how CSF-based molecular profiling can provide insight into biological determinants of neurological recovery. NPH represents a clinical context among causes of dementia in which substantial functional improvement can occur following a defined intervention, offering a unique opportunity to study recovery-associated neurobiology in humans. The metabolic programs implicated here, spanning one-carbon and redox metabolism and alternative substrate utilization, are increasingly recognized as central to neural resilience and neurological disease, suggesting that the biology identified in this setting may have broader relevance beyond NPH.^102–107^ Because Alzheimer’s disease-consistent CSF profiles and α-synuclein pathology can each be detected in roughly one-third of patients evaluated for iNPH,^108^ and shunt response appears less predictable in such complex presentations,^109^ profiling NPH alongside Alzheimer’s and Parkinson’s diseases and comorbid presentations is an important next step.

HVLP response is most commonly evaluated in terms of the gait symptom domain,^3,22–28,110^ whereas cognitive measures consistently fail to achieve equivalent discriminatory power at the pre-surgical stage,^111^ and Montreal Cognitive Assessment (MoCA) scores have been shown to not differ between surgical candidates and non-candidates even when objective gait measures do,^112^ partly because cognitive improvement depends on the time elapsed after lumbar puncture and the number of punctures performed.^113^ That the clearest metabolic association in our data concerned cognition suggests that CSF one-carbon metabolism is worth testing alongside HVLP. A comparison with HVLP is not possible here: the patients were selected for surgery partly on the basis of HVLP, their HVLP results were not available for analysis, and published HVLP accuracies refer mainly to gait or global response, use different outcome definitions and come from cohorts not selected on HVLP. Whether a metabolic marker adds to HVLP requires a head-to-head comparison in the same patients.

Several study limitations warrant consideration. The analytic cohort (n=86) excluded additional clinical subgroups, including patients evaluated but not selected for surgery, NPH patients who did not or could not consent to participate, and those with more granular outcome trajectories. As a single-center convenience sample comprising individuals approached by research staff who provided informed consent, the findings may not fully generalize to the broader NPH population. The cohort was predominantly White (87.2%), and differences across sociodemographic groups were not assessed. Outcomes were binary clinician judgements per domain, and patients who worsened were analyzed with those who did not improve. HVLP results, routine CSF laboratory values, comorbidities and the interval between lumbar and ventricular sampling were not available for analysis, and enrollment period was approximated by cohort. Because the sample sets partially overlapped and originated from a single center, the design provides internal verification rather than independent validation. The semi-targeted and transcriptomic data come from the same cohort 1 patients, and the transcriptomic data derive from CSF extracellular vesicles. A validation study estimating an AUC of 0.75 to within ±0.075 would need approximately 290 patients when sized for the cognitive outcome at its observed 21% non-improvement rate. For the gait and urinary outcomes, with higher non-improvement rates, roughly 200 patients would meet the same criterion (Supplementary Methods; Supplementary Table 3f; Supplementary Fig. 9).^85^

Outcomes were evaluated holistically by clinicians within each symptom domain at 3-month follow-up, consistent with the primary endpoint used in a recent randomized trial by Luciano et al.^4^ Although longer-term follow-up may have yielded different results, 85% of shunted patients returned for assessment, representing a strength of the study and likely reflecting the shorter follow-up interval. Finally, while metabolomic and transcriptional signals^43^ converged at the pathway level, with only 32 patients having both layers, we did not attempt multi-omics prediction models; integrating complementary molecular layers will require larger cohorts and biologically informed approaches.

Lumbar CSF is routinely collected during the diagnostic workup of NPH (HVLP) and is substantially more accessible than intraoperative ventricular CSF. Lumbar and ventricular CSF shared overall metabolite profiles, but in our 12 matched patients only the kynurenine/tryptophan ratio transferred closely between compartments, and plasma diverged substantially,^114^ so a targeted, quantitative lumbar CSF assay for the one-carbon/redox index with standardized pre-analytical handling will be needed.^115^ The locked index can then be evaluated in an independent, prospective cohort, reported according to TRIPOD+AI,^100^ including whether it provides incremental value beyond HVLP and existing biomarkers (Table 2).^97^

In summary, a CSF one-carbon/redox metabolic signature is associated with recovery after shunt surgery in NPH, most clearly for cognition, with convergent transcriptomic evidence from the same patients and a modest, prespecified-index-driven cognitive signal under strict cross-validation. These findings position one-carbon and choline metabolism as a candidate biological correlate of cognitive recovery to be tested prospectively.

## Data availability

The metabolomics data will be deposited in the Metabolomics Workbench (https://www.metabolomicsworkbench.org/), and the accession number will be provided on publication. For peer review, the data can be made available upon request to the corresponding authors.

The code for all analyses (locked dataset construction, indices, statistical models, nested cross-validation, permutation tests, transcriptomic integration and figures), together with the dated analysis log and the locked prediction settings, is publicly available at https://gitlab.com/nph_biomarkers. The code contains no patient-level data; input files are identified by their checksums.

## Acknowledgements

We thank Ryann Fame for insightful comments and critical review of the manuscript. We thank Shawn Kant, Tuan Pham, Thomas Serre, and Ritambhara Singh for their modeling insights. The clinical sample collection aspects of this study were approved as human subjects research by the Rhode Island Hospital institutional review board (#1492994), with written informed consent from all participants or their legally authorized representatives for sample collection and data abstraction from medical records. All human subjects research procedures were in accordance with the declaration of Helsinki and applicable local regulations.

We sincerely thank the patients and their families who participated in this study. Their generosity and willingness to contribute to research are essential for advancing the understanding and treatment of NPH and related neurological conditions.

## Author contributions

Conceptualization: A.F., M.G.R., B.P.

Methodology: B.P., L.D., J.R.B., R.M., A.F., M.G.R.

Investigation: B.P., L.D., J.R.B., R.M., M.T., O.P.L., A.H., S.S., K.A.S., M.G.R., N.A.-N., S.A.

Formal analysis: B.P., L.D., C.P.S., J.R.B., R.M.

Data curation: B.P., L.D., J.R.B., R.M., M.T., O.P.L., A.H., S.S.

Visualization: B.P., L.D., R.M.

Resources: M.T., O.P.L., A.H., S.S., P.M.K., K.A.S., M.G.R., N.A.-N., S.A.

Supervision: A.F., M.G.R., N.K., B.P.

Writing – original draft: M.G.R., L.D., A.F., B.P.

Writing – review & editing: P.M.K., O.P.L., M.T., A.H., C.P.S., A.F., M.G.R., B.P., and all authors.

Project administration: B.P., A.F., N.K., M.G.R. Funding acquisition: A.F., N.K., M.G.R.

## Supplementary material

Supplementary material is available at Brain online: Supplementary Figs 1–9 with legends, Supplementary Methods, Supplementary Tables 1–5 and Supplementary Data 1–2. Supplementary Data 1, the 455 metabolite annotations of the semi-targeted platform; Supplementary Data 2, the metabolites of the targeted platform; Supplementary Table 1, prespecified index tests with sensitivity and per-domain analyses; Supplementary Table 2, per-metabolite analysis; Supplementary Table 3, prediction models (nested cross-validation, permutation tests, calibration and classification metrics, incremental value, the estimates of the initial exploratory pipeline and its nested leave-one-out re-analysis, precision and validation sample size); Supplementary Table 4, transcriptomic integration (enzyme–metabolite pairs and gene-set tests); Supplementary Table 5, ventricular–lumbar concordance. De-identified patient-level clinical and metabolomic data can be made available upon request (see Data availability).

