## Supplementary material for "A cerebrospinal fluid metabolic signature of recovery in normal pressure hydrocephalus"

***Running title**: CSF metabolomics and recovery in NPH*

Authors: Lisa J. Duan[1], Mei E. Tiemeyer[2], Owen P. Leary[2,3], Amanda Hasbrouck[2], Caleb P. Schultz[4], Rohan Meier[1], Shanzeh Sayied[2,5], Natalie Amaral-Nieves[2,3], Jeannette Riley Brook[5], Naama Kanarek[6,7,8], Saud Alushaini[9], Maria Guglielmo[2,3,10], Konstantina A. Svokos[2,3,10], Petra M. Klinge[2,3,10], Alexander Fleischmann[1,10], Maria Grazia Ruocco[4,11,#], Boryana Petrova[12, #]

[1] Department of Neuroscience, Brown University, RI, USA

[2] Department of Neurosurgery, Rhode Island Hospital, RI, USA

[3] The Warren Alpert Medical School of Brown University, RI, USA

[4] Adelle Diagnostics Inc, 225 Dyer St, 2nd Floor Providence, RI 02903

[5] Albert Einstein College of Medicine, New York, NY, USA

[6] Department of Pathology, Boston Children's Hospital, Boston, MA 02115, USA

[7] Harvard Medical School, Boston, MA 02115, USA

[8] Broad Institute of MIT & Harvard, 415 Main St, Cambridge, MA 02142, USA

[9] Department of Neurology, Rhode Island Hospital, RI, USA

[10] Carney Institute for Brain Science, Brown University, RI, USA

[11] Department of Cognitive & Psychological Sciences, Brown University, RI, USA

[12] Medical University of Vienna, Vienna 1090, Austria

### Supplementary Figures and Figure Legends

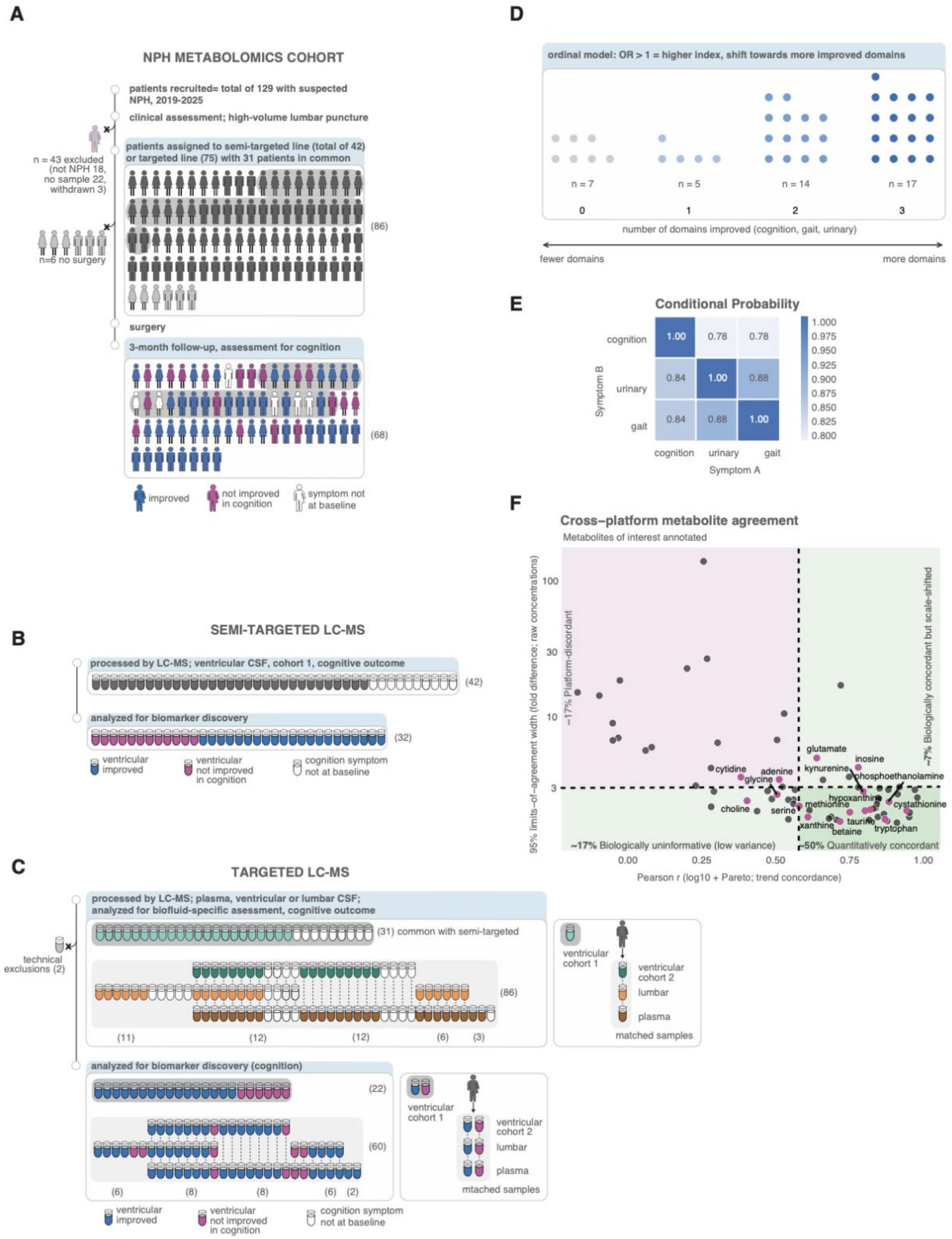

**Supplementary Fig. 1. NPH cohort composition and sample allocation across analytical stages. (A)** Overview of the patient cohort. Of 129 patients recruited with suspected NPH (2020–2025), 43 were excluded (not NPH, 18; no biofluid sample, 22; withdrawn, 3), leaving 86 NPH patients with at least one biofluid sample (numbers in parentheses). Upper grid: one icon per patient, shown as female or male, assigned to the semi-targeted line only (n = 11), both lines (grey shading, n = 31) or the targeted line only (n = 44); semi-targeted line, 42 patients in total; targeted line, 75. Six patients (all targeted line only; 3 female, 3 male) did not undergo surgery (light grey). Lower grid: the 68 shunted patients with a 3-month assessment, in the same order, coloured by cognitive outcome as a representative clinical outcome measure (improved, n = 44; not improved, n = 18; cognitive symptom absent at baseline, n = 6). The 12 shunted patients without a 3-month assessment and the 6 without surgery are not shown. Semi-targeted line, cognitive outcome: 20 improved (4 unique, 16 in both lines), 12 not improved (6 unique, 6 in both), 6 symptom absent (1 unique, 5 in both) and 4 without 3-month assessment (all in both). Targeted line, cognitive outcome: 40 improved (24 unique, 16 in both), 12 not improved (6 unique, 6 in both), 5 symptom absent (all in both), 12 without 3-month assessment (8 unique, 4 in both) and 6 without surgery (all unique). (**B**) Semi-targeted LC–MS: ventricular CSF of 42 cohort-1 patients was processed; 32 had a cognitive outcome and were analyzed for biomarker discovery (20 improved, 12 not improved). Of the remaining 10, 6 had no cognitive symptom at baseline and 4 had no 3-month assessment. (**C**) Targeted LC–MS: plasma, ventricular and lumbar CSF samples, comprising samples re-measured from the semi-targeted cohort and newly collected samples (numbers in parentheses, sample counts). Two ventricular samples were excluded for technical reasons (one not acquired, one failed predefined quality control). Grey box: the 31 ventricular CSF samples in common with the semi-targeted cohort (ventricular cohort 1); cognitive outcome 16 improved, 6 not improved, 5 symptom absent at baseline and 4 without 3-month assessment. The 86 cohort-2 ventricular, lumbar and plasma samples are shown with their matched sets: lumbar CSF only (n = 11; 4 improved, 2 not improved, 1 without assessment, 4 without surgery), ventricular cohort 2 with lumbar CSF and plasma (n = 12; 7 improved, 1 not improved, 4 without assessment), ventricular cohort 2 with plasma (n = 12; 7 improved, 1 not improved, 3 without assessment, 1 without surgery), lumbar CSF with plasma (n = 6; 4 improved, 2 not improved), and plasma only (n = 3; 2 improved, 1 without surgery). Lower box: samples with a cognitive outcome, analyzed for biomarker discovery and biofluid comparison (ventricular cohort 1, n = 22; cohort 2 and other fluids, n = 60). Improved, blue; not improved in cognition, magenta; cognitive symptom absent at baseline, white. (**D**) Distribution of the primary outcome, the number of domains improved (cognition, gait, urinary; 0–3) among patients with all baseline-symptomatic domains scored (n = 43: 7, 5, 14 and 17 patients with 0, 1, 2 and 3 domains improved). In the proportional-odds model, an odds ratio above 1 indicates that a higher index value shifts patients towards more improved domains. (**E**) Conditional probability of improvement in one domain (symptom B) given improvement in another (symptom A), among patients symptomatic in both; blue gradient. (**F**) Cross-platform agreement for metabolites measured on both the semi-targeted and targeted platforms in overlapping samples. x-axis, Pearson correlation of log~10~-transformed, Pareto-scaled values (trend concordance); y-axis, width of the Bland–Altman 95% limits of agreement on raw values (fold difference). Dashed lines, [r = 60] and 3-fold. Quadrants: quantitatively concordant (~50%), biologically concordant but scale-shifted (~7%), biologically uninformative with low variance (~17%) and platform-discordant (~17%). Metabolites of interest are labelled.

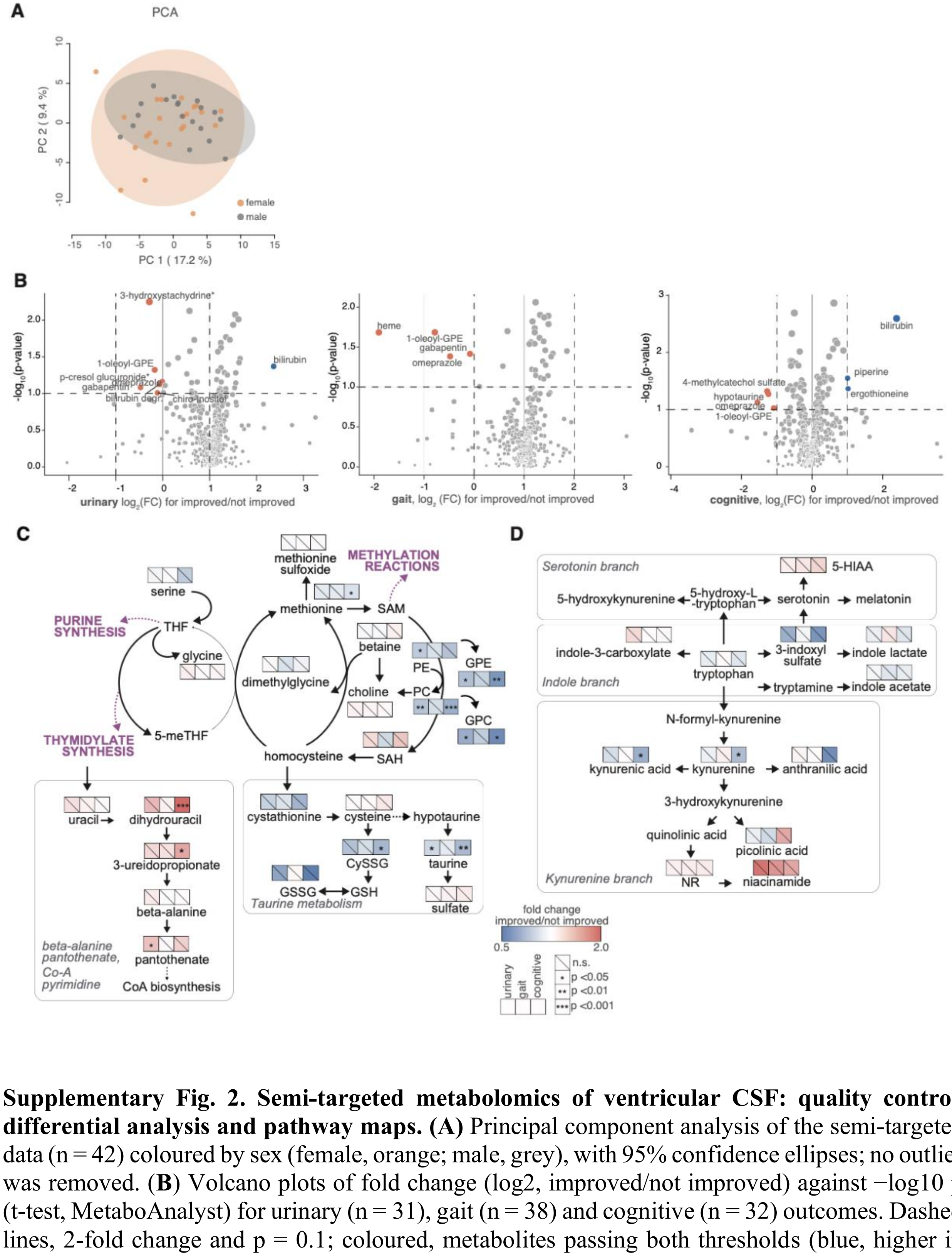


**Supplementary Fig. 2. Semi-targeted metabolomics of ventricular CSF: quality control, differential analysis and pathway maps. (A)** Principal component analysis of the semi-targeted data (n = 42) coloured by sex (female, orange; male, grey), with 95% confidence ellipses; no outlier was removed. (**B**) Volcano plots of fold change (log2, improved/not improved) against −log10 p (t-test, MetaboAnalyst) for urinary (n = 31), gait (n = 38) and cognitive (n = 32) outcomes. Dashed lines, 2-fold change and p = 0.1; coloured, metabolites passing both thresholds (blue, higher in improved; orange, lower in improved). None passed FDR < 0.05. (**C**) Semi-targeted pathway map of one-carbon metabolism and its exits: methylation and the choline branch (choline, phosphocholine, glycerophosphocholine, phosphoethanolamine, glycerophosphoethanolamine), transsulfuration and taurine metabolism, and thymidylate synthesis leading to pyrimidine catabolism, β-alanine and CoA. Squares as in Fig. 5A (fold change improved/not improved; stars, p from covariate-adjusted per-metabolite models, age and sex). CySSG, cysteine-glutathione disulfide; GPC, glycerophosphocholine; GPE, glycerophosphoethanolamine. (**D**) As in (**C**), for tryptophan metabolism. Quinolinate and hypotaurine were detected in fewer than half of the samples and were not tested; nodes without squares were not measured.

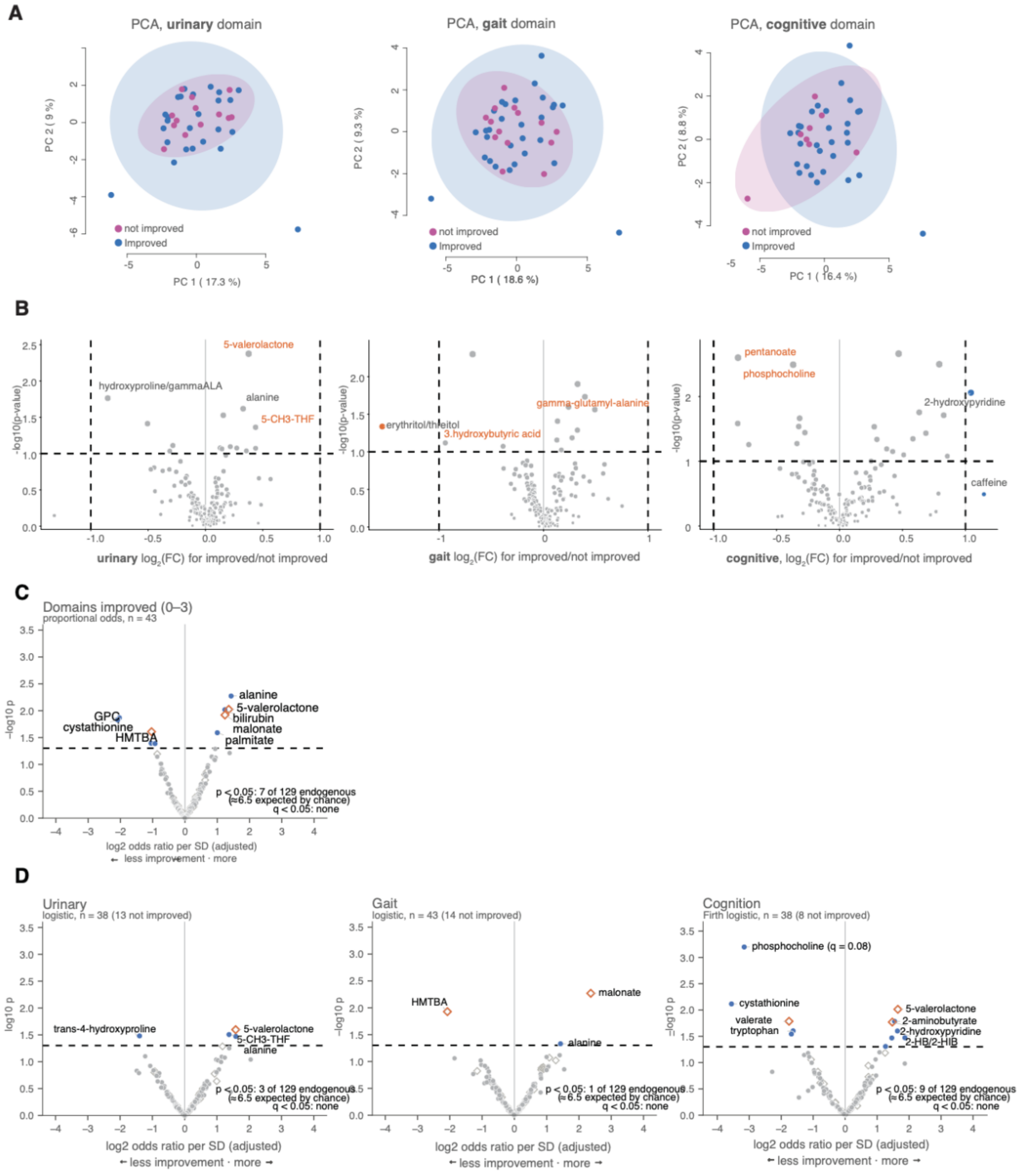


**Supplementary Fig. 3. Targeted metabolomics of ventricular CSF reveals pathways associated with urinary, gait, and cognitive improvement after NPH surgery.** (**A**) Principal component analysis per domain (urinary n = 38, gait n = 43, cognition n = 38), coloured by outcome, with 95% confidence ellipses; no sample was excluded. (**B**) Fold-change volcano plots (log2, improved/not improved; t-test, MetaboAnalyst) per domain; dashed lines, 2-fold change and

$p = 0.1$; metabolites of interest in orange. None passed FDR $< 0.05$. (**C**) Covariate-adjusted per-metabolite analysis (Supplementary Table 2): each point is one targeted metabolite, adjusted for age, sex and enrolment cohort; x, log2 odds ratio per SD (positive, more improvement); y, −log10 p. Number of domains improved (proportional odds, $n = 43$), urinary and gait (logistic regression) and cognition (Firth-penalized logistic regression). Filled circles, endogenous metabolites (one Benjamini–Hochberg family per outcome); open diamonds, microbial, dietary, drug-derived or uncertain origin (separate family); coloured, $p < 0.05$; dashed line, $p = 0.05$; q shown where $< 0.10$. Number of endogenous metabolites with $p < 0.05$ out of 129: domains 7, urinary 3, gait 1, cognition 9 (6.5 expected by chance); none with $q < 0.05$.

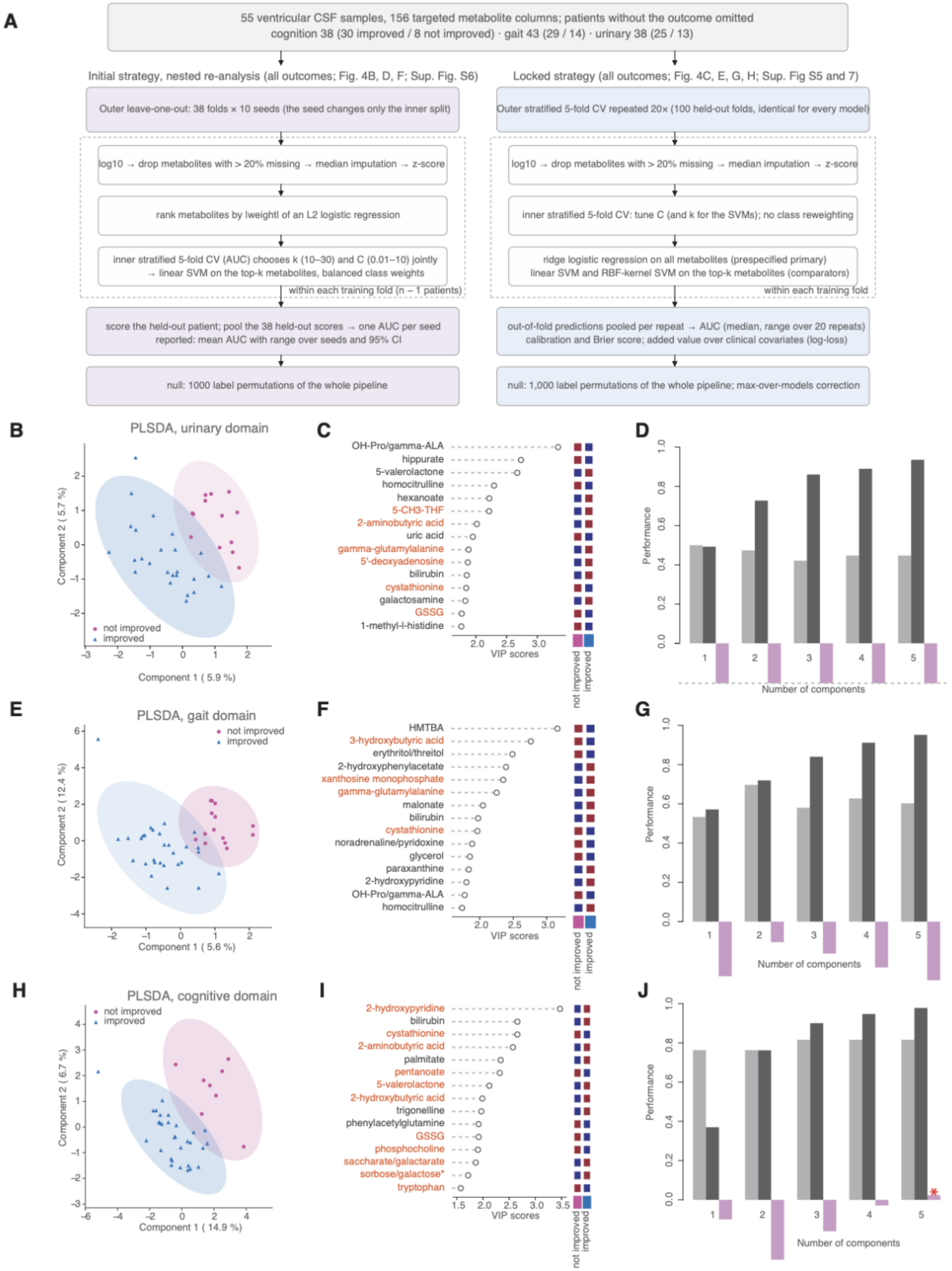

A
55 ventricular CSF samples, 156 targeted metabolite columns; patients without the outcome omitted
cognition 38 (30 improved / 8 not improved) · gait 43 (29 / 14) · urinary 38 (25 / 13)
Initial strategy, nested re-analysis (all outcomes; Fig. 4B, D, F; Sup. Fig. S6)
Outer leave-one-out: 38 folds × 10 seeds (the seed changes only the inner split)
log10 → drop metabolites with > 20% missing → median imputation → z-score
rank metabolites by |weight| of an L2 logistic regression
inner stratified 5-fold CV (AUC) chooses k (10–30) and C (0.01–10) jointly
→ linear SVM on the top-k metabolites, balanced class weights
within each training fold (n − 1 patients)
score the held-out patient; pool the 38 held-out scores → one AUC per seed
reported: mean AUC with range over seeds and 95% CI
null: 1000 label permutations of the whole pipeline
Locked strategy (all outcomes; Fig. 4C, E, G, H; Sup. Fig S5 and 7)
Outer stratified 5-fold CV repeated 20× (100 held-out folds, identical for every model)
log10 → drop metabolites with > 20% missing → median imputation → z-score
inner stratified 5-fold CV: tune C (and k for the SVMs); no class reweighting
ridge logistic regression on all metabolites (prespecified primary)
linear SVM and RBF-kernel SVM on the top-k metabolites (comparators)
within each training fold
out-of-fold predictions pooled per repeat → AUC (median, range over 20 repeats)
calibration and Brier score; added value over clinical covariates (log-loss)
null: 1,000 label permutations of the whole pipeline; max-over-models correction
B
PLSDA, urinary domain
Component 1 ( 5.9 %)
Component 2 ( 5.7 %)
not improved
improved
C
OH-Pro/gamma-ALA
hippurate
5-valerolactone
homocitrulline
hexanoate
5-CH3-THF
2-aminobutyric acid
uric acid
gamma-glutamylalanine
5'-deoxyadenosine
bilirubin
cystathionine
galactosamine
GSSG
1-methyl-l-histidine
VIP scores
not improved
improved
D
Performance
Number of components
E
PLSDA, gait domain
Component 1 ( 5.6 %)
Component 2 ( 12.4 %)
not improved
improved
F
HMTBA
3-hydroxybutyric acid
erythritol/threitol
2-hydroxyphenylacetate
xanthosine monophosphate
gamma-glutamylalanine
malonate
bilirubin
cystathionine
noradrenaline/pyridoxine
glycerol
paraxanthine
2-hydroxypyridine
OH-Pro/gamma-ALA
homocitrulline
VIP scores
not improved
improved
G
Performance
Number of components
H
PLSDA, cognitive domain
Component 1 ( 14.9 %)
Component 2 ( 6.7 %)
not improved
improved
I
2-hydroxypyridine
bilirubin
cystathionine
2-aminobutyric acid
palmitate
pentanoate
5-valerolactone
2-hydroxybutyric acid
trigonelline
phenylacetylglutamine
GSSG
phosphocholine
saccharate/galactarate
sorbose/galactose*
tryptophan
VIP scores
not improved
improved
J
Performance
Number of components
*

**Supplementary Fig. 4. Prediction strategies and PLS-DA of targeted ventricular CSF metabolomics.** (**A) The two nested** prediction **strategies. Left, the leave-one-out pipeline of the initial analysis, re-run in nested form after the analysis lock for all three outcomes (cognition, Fig. 4B, D, F; gait and urinary, Supplementary Fig. 6 and Supplementary Table 3d). Right, the locked** protocol **for all outcomes (ML1–ML4; Fig. 4C, E, G, H and Supplementary Figs 5 and 7).** Of 55 ventricular CSF samples (156 targeted metabolite columns), patients without an outcome were omitted (urinary 17, gait 12, cognition 17), leaving cognition 38 (30 / 8), gait 43 (29 / 14) and urinary 38 (25 / 13). **Locked protocol: outer** stratified 5-fold cross-validation repeated 20 times (100 held-out folds, identical for every model); within each training fold: log10 transformation, removal of metabolites with >20% missing values, median imputation, z-scoring and an inner 5-fold cross-validation to tune the penalty C (and the number of metabolites k for the SVMs). Primary model, ridge logistic regression on all metabolites; comparators, linear and RBF-kernel SVMs on the top-k metabolites ranked by logistic-regression weight. Evaluation: out-of-fold predictions pooled over repeats; discrimination and calibration (ML2); permutation null of the whole pipeline (1,000 label shuffles, 5 repeats each) with max-over-models correction (ML3); added value over clinical covariates with indices recomputed inside each training fold, log-loss as the primary comparison (ML4). (**B–J**) PLS-DA (MetaboAnalyst; targeted data, log-transformed and Pareto-scaled) for urinary (**B–D**), gait (**E–G**) and cognition (**H–J**). (B, E, H) Scores on components 1 and 2 (explained variance on the axes) with 95% confidence ellipses; not improved, magenta circles; improved, blue triangles. (**C, F, I**) Top 15 metabolites by variable importance in projection (VIP, component 1); orange, nominal $p < 0.05$ in the covariate-adjusted per-metabolite analysis for the same outcome; squares, higher (red) or lower (blue) group mean of autoscaled log10 values in not improved and improved patients. (**D, G, J**) 5-fold cross-validated accuracy, $R^2$ and $Q^2$ by number of components; $Q^2$ below zero indicates that cross-validated predictions are worse than predicting the group mean, i.e. the PLS-DA models fit the data (high $R^2$) but do not generalize; asterisk, number of components selected by MetaboAnalyst; negative $Q^2$ values are truncated at −0.2 for clarity (dashed line). PLS-DA permutation test: $p = 0.32$ (urinary), 0.35 (gait) and 1.00 (cognition).

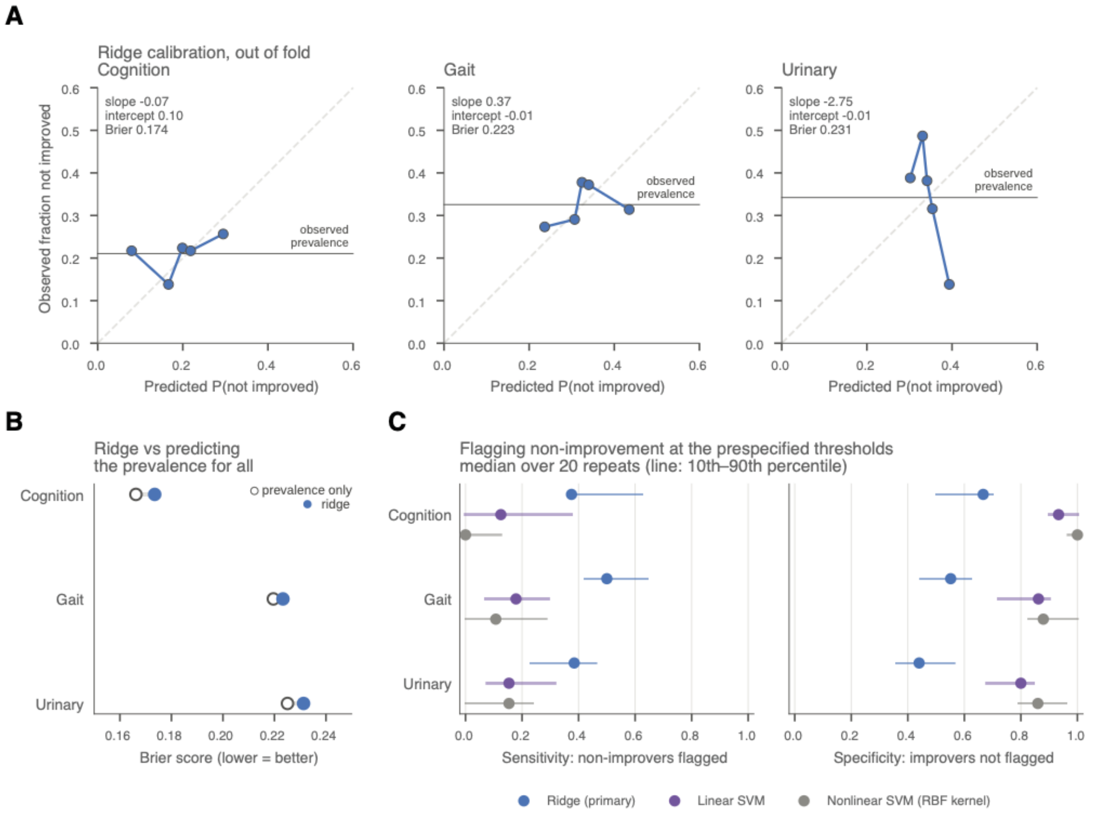


**Supplementary Fig. 5. Calibration and classification of the prediction models. (A)** Calibration of the ridge model (primary) for cognition, gait and urinary outcomes: out-of-fold predicted probability of non-improvement, pooled over the 20 repeats and binned into quintiles, against the observed fraction not improved; dashed diagonal, perfect calibration; horizontal line, observed prevalence. Calibration slope, intercept and Brier score per outcome (cognition −0.07, 0.10, 0.173; gait 0.37, −0.01, 0.223; urinary −2.75, −0.01, 0.231). (**B**) Brier score of the ridge model (filled) against a model that predicts the prevalence for everyone (open; lower is better). (**C**) Sensitivity (non-improvers flagged) and specificity (improvers not flagged) at the thresholds fixed in advance (ridge, predicted probability of improvement below the prevalence; SVMs, decision score below zero); points, median over 20 repeats; lines, 10th–90th percentile. Ridge (primary), blue; linear SVM, purple; RBF-kernel SVM, grey.

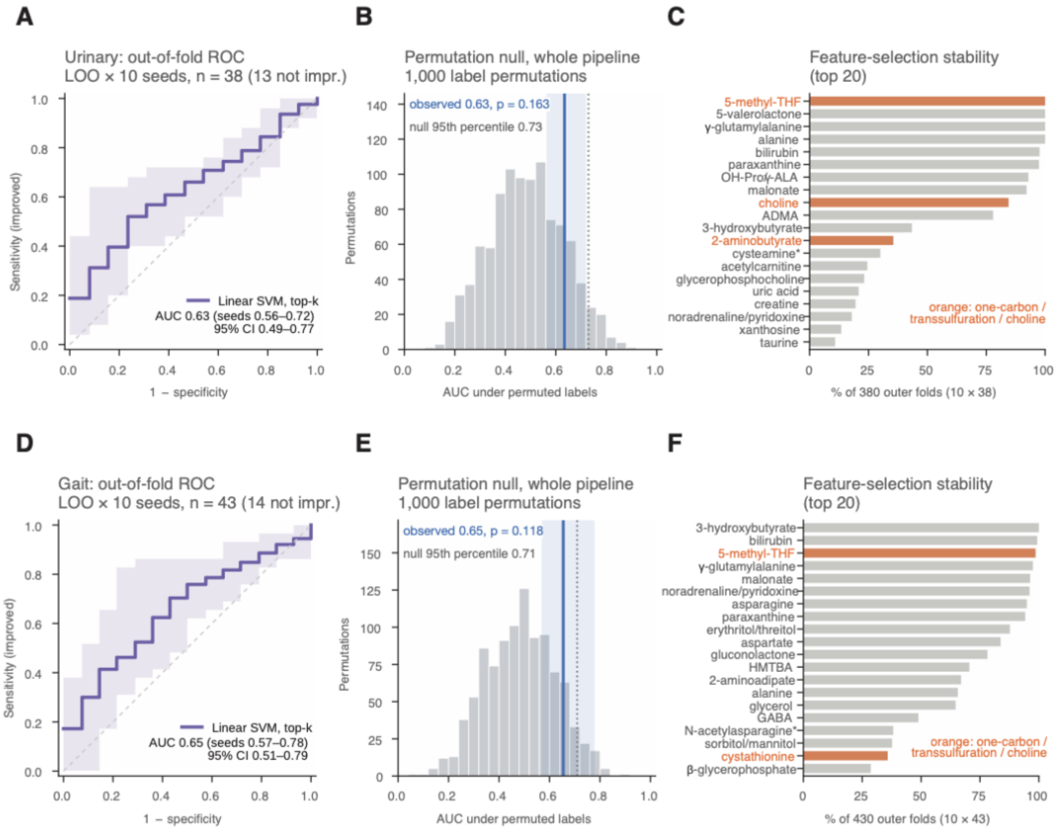


**Supplementary Fig. 6.** Leave-one-out pipeline for urinary and gait outcomes. As **Fig. 4B, D** and F, for urinary (**A–C**; n = 38, 13 not improved) and gait (**D–F**; n = 43, 14 not improved) outcomes: the leave-one-out pipeline of the initial analysis re-run in nested form after the analysis lock (10 seeds, which change only the inner split; preprocessing as in the initial analysis: log2 transformation and, within each training fold, removal of metabolites with >20% missing values, mean imputation and autoscaling). (**A, D**) Out-of-fold ROC curve of the linear SVM on the top-k metabolites; mean curve across seeds, shaded band, range across seeds; dashed diagonal, chance. Mean AUC (range across seeds; 95% patient-bootstrap CI): urinary 0.63 (0.56–0.72; 0.49–0.77); gait 0.65 (0.57–0.78; 0.51–0.79). (**B, E**) Permutation null of the whole pipeline: AUC distribution from 1,000 label permutations (grey), null 95th percentile (dotted), observed mean AUC (line) and range of the observed AUC across seeds (shaded box); urinary, null 95th percentile 0.73, p = 0.16; gait, 0.71, p = 0.12. (**C, F**) Feature-selection stability: percentage of outer folds (10 seeds × number of patients) in which each metabolite was selected (top 20); orange, one-carbon, transsulfuration and choline metabolites. Neither outcome exceeded its permutation null.

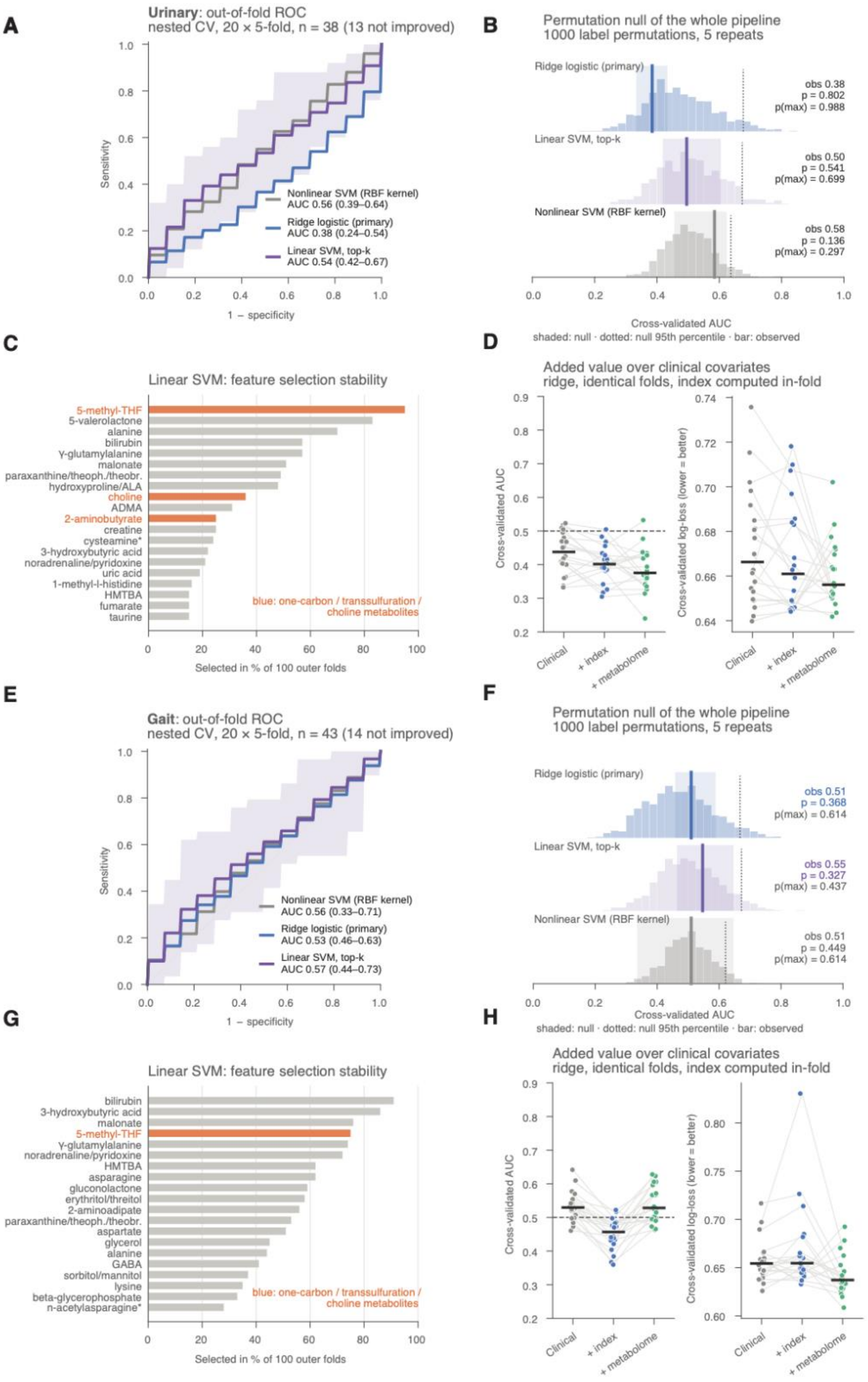
A
Urinary: out-of-fold ROC
nested CV, 20 × 5-fold, n = 38 (13 not improved)
Sensitivity
1 − specificity
Nonlinear SVM (RBF kernel)
AUC 0.56 (0.39–0.64)
Ridge logistic (primary)
AUC 0.38 (0.24–0.54)
Linear SVM, top-k
AUC 0.54 (0.42–0.67)
B
Permutation null of the whole pipeline
1000 label permutations, 5 repeats
Ridge logistic (primary)
obs 0.38
p = 0.802
p(max) = 0.988
Linear SVM, top-k
obs 0.50
p = 0.541
p(max) = 0.699
Nonlinear SVM (RBF kernel)
obs 0.58
p = 0.136
p(max) = 0.297
Cross-validated AUC
shaded: null · dotted: null 95th percentile · bar: observed
C
Linear SVM: feature selection stability
5-methyl-THF
5-valerolactone
alanine
bilirubin
γ-glutamylalanine
malonate
paraxanthine/theoph./theobr.
hydroxyproline/ALA
choline
ADMA
2-aminobutyrate
creatine
cysteamine*
3-hydroxybutyric acid
noradrenaline/pyridoxine
uric acid
1-methyl-l-histidine
HMTBA
fumarate
taurine
blue: one-carbon / transsulfuration / choline metabolites
Selected in % of 100 outer folds
D
Added value over clinical covariates
ridge, identical folds, index computed in-fold
Cross-validated AUC
Cross-validated log-loss (lower = better)
Clinical
+ index
+ metabolome
E
Gait: out-of-fold ROC
nested CV, 20 × 5-fold, n = 43 (14 not improved)
Sensitivity
1 − specificity
Nonlinear SVM (RBF kernel)
AUC 0.56 (0.33–0.71)
Ridge logistic (primary)
AUC 0.53 (0.46–0.63)
Linear SVM, top-k
AUC 0.57 (0.44–0.73)
F
Permutation null of the whole pipeline
1000 label permutations, 5 repeats
Ridge logistic (primary)
obs 0.51
p = 0.368
p(max) = 0.614
Linear SVM, top-k
obs 0.55
p = 0.327
p(max) = 0.437
Nonlinear SVM (RBF kernel)
obs 0.51
p = 0.449
p(max) = 0.614
Cross-validated AUC
shaded: null · dotted: null 95th percentile · bar: observed
G
Linear SVM: feature selection stability
bilirubin
3-hydroxybutyric acid
malonate
5-methyl-THF
γ-glutamylalanine
noradrenaline/pyridoxine
HMTBA
asparagine
gluconolactone
erythritol/threitol
2-aminoadipate
paraxanthine/theoph./theobr.
aspartate
glycerol
alanine
GABA
sorbitol/mannitol
lysine
beta-glycerophosphate
n-acetylasparagine*
blue: one-carbon / transsulfuration / choline metabolites
Selected in % of 100 outer folds
H
Added value over clinical covariates
ridge, identical folds, index computed in-fold
Cross-validated AUC
Cross-validated log-loss (lower = better)
Clinical
+ index
+ metabolome

**Supplementary Fig. 7. Prediction of urinary and gait improvement.** As the locked-protocol panels of Fig. 4 (**C, E, G, H**), for urinary (A–D; n = 38, 13 not improved) and gait (**E–H**; n = 43, 14 not improved) outcomes. (**A, E**) Out-of-fold ROC curves; median AUC (range across repeats): urinary, ridge 0.38 (0.24–0.54), linear SVM 0.54 (0.42–0.67), RBF-kernel SVM 0.56 (0.39–0.64); gait, ridge 0.53 (0.46–0.63), linear SVM 0.57 (0.44–0.73), RBF-kernel SVM 0.56 (0.33–0.71). (**B, F**) Permutation null (1,000 permutations, 5 repeats; box, range of the observed AUC across the 5 repeats): observed AUC, p and p(max): urinary, ridge 0.38, 0.802, 0.988; linear SVM 0.50, 0.541, 0.699; RBF-kernel SVM 0.58, 0.136, 0.297; gait, ridge 0.51, 0.368, 0.614; linear SVM 0.55, 0.327, 0.437; RBF-kernel SVM 0.51, 0.449, 0.614. (**C, G**) Selection frequency in the linear SVM across 100 outer folds (top 20); orange, one-carbon, transsulfuration and choline metabolites. (**D, H**) Incremental value over clinical covariates (AUC, left; log-loss, right); neither the indices nor the metabolome improved on clinical covariates.

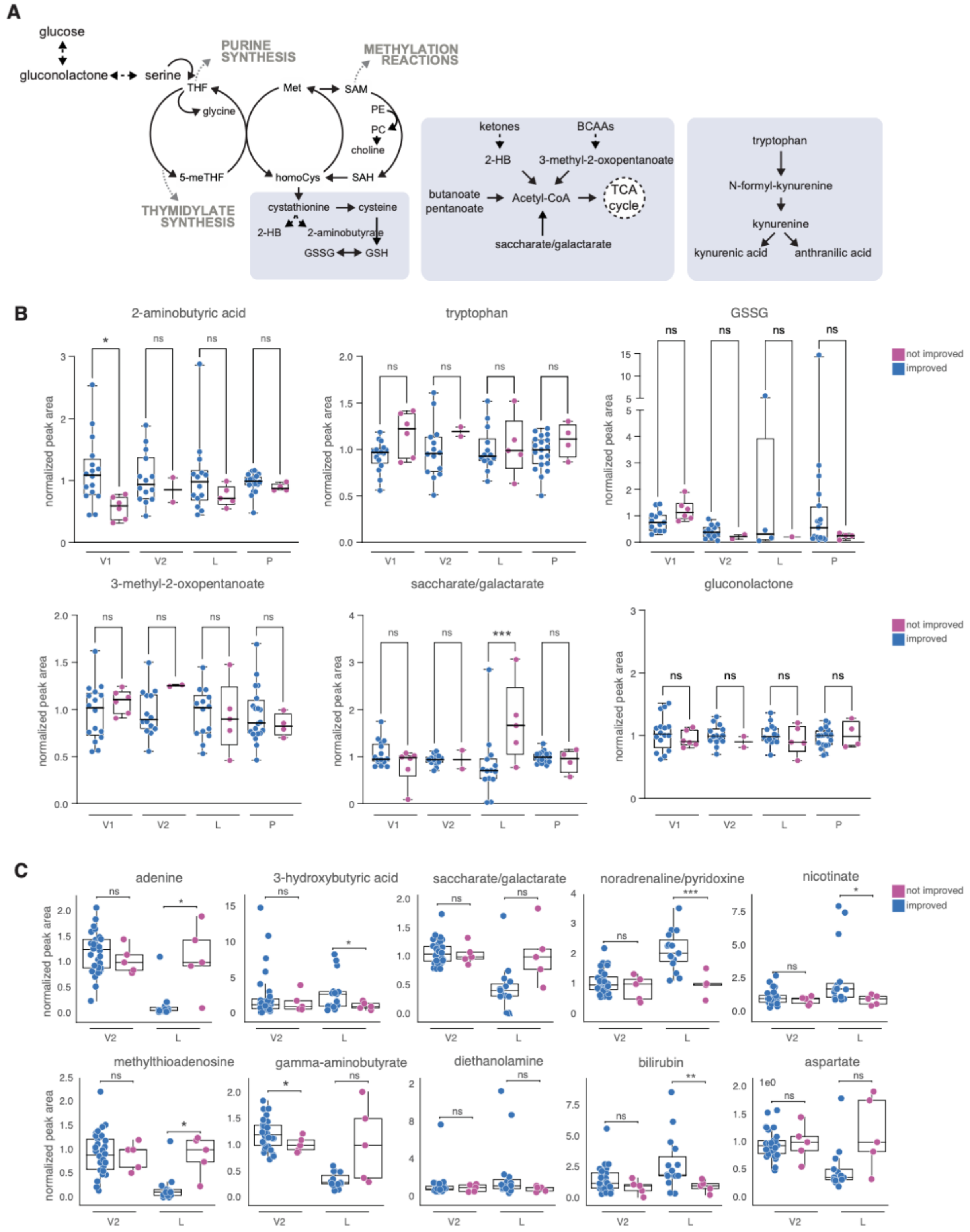


**Supplementary Fig. 8. Recovery-associated metabolites across biofluids (cognition)**. (A) Pathway context of the metabolites shown (one-carbon and transsulfuration; alternative carbon sources; kynurenine branch). (**B**) Selected metabolites in ventricular CSF cohort 1 (V1), ventricular CSF cohort 2 (V2), lumbar CSF (L) and plasma (P), by cognitive outcome (improved, blue; not improved, magenta), expressed as fold of the mean of all locked samples of that fluid; boxes, median and interquartile range; whiskers, **minimum to maximum.** One-way ANOVA with Šídák's multiple comparisons test (ns, not significant; *p = 0.01; ***p = 0.0001; ****p < 0.0001). Cognition n: V1 22 (16 / 6), V2 16 (14 / 2), L 20 (15 / 5), P 24 (20 / 4). (**C**) Metabolites differing between outcome groups in lumbar CSF, shown for V2 and L. Because of the small number of non-improved patients in each fluid (2–6), these comparisons are descriptive.

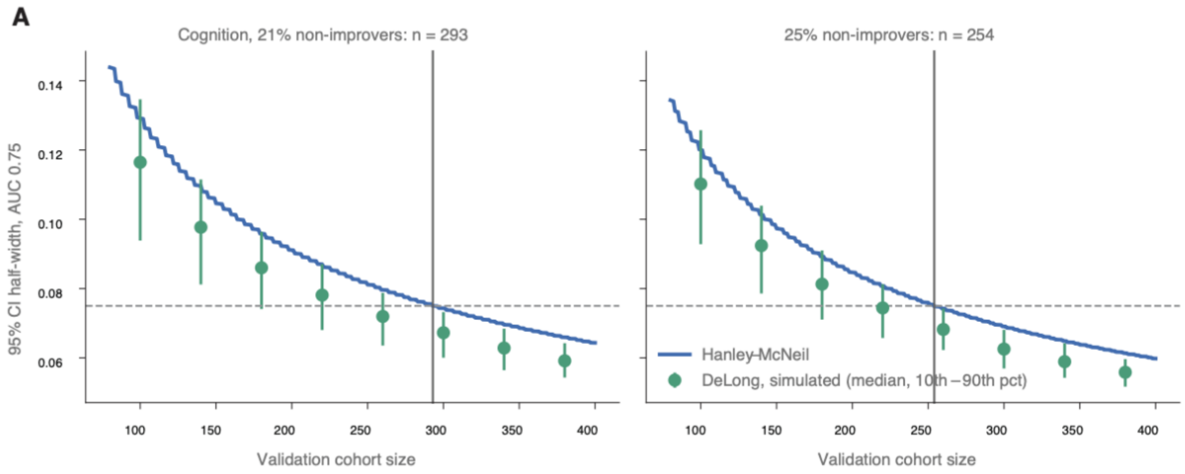


**Supplementary Fig. 9.** (**A**)Validation sample size. 95% confidence half-width of an AUC of 0.75 against validation cohort size, from the Hanley–McNeil standard error (line) and from DeLong intervals in 500 simulated[1] cohorts per size (points, median; bars, 10th–90th percentile), at the observed cognitive non-improvement rate (21%; left) and at 25% (right). Dashed line, target half-width ±0.075; vertical line, required size (293 and 254 patients).

### Supplementary Methods

*Patient cohort*

Research samples included in analyses (lumbar CSF, ventricular CSF and plasma samples) were collected from 86 patients evaluated for suspected normal pressure hydrocephalus (NPH) at Rhode Island Hospital/Brown University Health between 1/2020 and 4/2025. The demographics of the analytic cohort are described in **Table 1**. This cohort represents a subset of all patients who were seen in either single-provider neurosurgical clinics (P.M.K. & K.S., 2019–2021) or a multi-provider multidisciplinary NPH clinic (2022–2025). From 2022 onward, the clinic where patients were diagnosed operated as a multidisciplinary program comprising neurologists, neurosurgeons, physical therapists, and neuropsychologists.[2] All patients were initially referred for suspected NPH and underwent standardized neurological examination at each visit. Clinician- and patient-reported outcomes were prospectively recorded in the electronic medical record and subsequently abstracted for research analysis. All patients were initially referred for suspected NPH and provided consent for research sample donation during routine procedures.

A total of n=129 patients provided informed consent, of whom n=86 were diagnosed with NPH, did not withdraw participation during the follow-up period, and had at least one sample collected by research staff during qualifying procedures. A total of 43 patients were excluded from the final analytic cohort for those three reasons (n=18, n=3, and n=22, respectively). Of the 43 excluded, samples were not ultimately collected and/or analyzed from 33, while the remaining 10 had samples collected and analyzed for metabolomics, but were excluded from final analyses due to not ultimately being diagnosed with NPH after full diagnostic work-up. The study protocol changed partway through the project period to include lumbar CSF and plasma samples in addition to ventricular CSF samples, and furthermore not all patients underwent diagnostic HVLP. Hence, there are fewer lumbar CSF and plasma samples (versus ventricular CSF samples) in the analytic dataset. During the study period, samples from consented, otherwise qualified participants with suspected or diagnosed NPH were occasionally missed due to staff availability (particularly during the COVID-19 pandemic lockdowns) and other logistical barriers (e.g., inability to collect sufficient sample during HVLP).

Clinical data collection and assessment procedures followed the protocol previously described in Levin et al.[3] In brief, clinical outcomes before and after shunt surgery were evaluated across the three components of the NPH triad - urinary incontinence, gait, and cognition. Symptoms were evaluated as present or absent at baseline and documented along with details of functional impact on activities of daily living and patient-reported evaluations consistent with prior NPH outcome reporting approaches.[4] Each domain was subsequently classified as improved or not improved after surgery by comparing follow-up clinic note documentation with pre-operative clinic note documentation by members of the research team who were blinded to metabolomic analyses. Improvement at 3-month follow-up was defined as follows: (1) Urinary improvement: reduction in frequency or resolution of urinary urgency or incontinence episodes based on patient report and clinician assessment. (2) Gait improvement: functional improvement in ambulation, including transition from walker to cane or independent walking, improved balance or stride length, improved walking speed or foot clearance, or reduction in frequency of falls. (3) Cognitive improvement: patients were considered improved if the patient and/or family member reported relevant changes in the quality of at least one of the following symptoms: improved forgetfulness; improved thought processing; increase in engaged conversations; resolution of lethargy and depression.

Outcomes were independently evaluated by neurosurgeons (P.M.K., K.S., M.G.) and recorded as part of routine clinical documentation. Data were entered into the electronic medical record as part of standard clinical documentation and later abstracted by study team members (O.P.L., M.T., S.S., and A. H.) for use as study endpoints.

All participants were informed of the study's purpose and provided written informed consent before participation, including consent to publish results in an anonymous format. The study was approved by the Institutional Review Board at Rhode Island Hospital (Brown University Health), IRB number: 1492994.

*Clinical outcome association analyses*

To determine whether improvements across clinical symptom domains (urinary, gait, cognitive) were interrelated following shunt surgery, each domain was binarized as “improved” or “not improved” according to predefined clinical criteria. For each pair of domains, conditional probabilities of improvement (e.g., P[urinary | gait]) were calculated. The strength and significance of associations were further evaluated using Pearson’s chi-square test and corresponding odds ratios, implemented with scipy.stats.chi2_contingency in Python 3.9.16. Results were visualized using bar plots generated with matplotlib.

*Biofluid sample collection*

Patient ventricular CSF samples were collected intraoperatively during ventriculoperitoneal shunt placement procedures. No duplicate samples of a given sample type were collected from the same individual. Each 15 ml sample was procured immediately upon placement of the ventricular catheter into a sterile, chilled polypropylene collection tube. Lumbar CSF samples were collected at outpatient diagnostic lumbar puncture procedures. Any CSF drained during these procedures, and not being used for clinical lab testing, was allocated to sterile polypropylene collection tubes for immediate transfer to the research team. Following both lumbar and ventricular CSF collections, 150 µl of samples was transferred to a micro cryotube (Azenta Life Sciences, 67-0753-11 FluidX™, Burlington, MA, USA), labeled with subject ID, spun at 4º C, flash frozen in liquid nitrogen, and stored at −80 °C until analysis. The remaining CSF was divided into 0.4 mL aliquots labeled with subject ID and “B” indicating backup, flash frozen in liquid nitrogen, and stored at −80 °C until analysis.

5-6 ml of whole blood was collected by venipuncture into EDTA-coated collection tubes (Cat/Ref#: 456038, Grenier Bio-One GmbH, Bad Haller Str. 32 4550 Kremsmunster, Austria). Blood was centrifuged at 1,300 x g at 4°C for 10 minutes. Resulting plasma was transferred into a polypropylene tube and 150 ul aliquots in micro cryotubes were flash frozen in liquid nitrogen, and stored at -80 °C until analysis.

*Semi-targeted metabolomics data collection*

A total of 60 µL CSF per patient was analyzed using the Precision Metabolomics platform from Metabolon Inc (Durham, NC, USA). Samples were processed under a standardized workflow that integrates multiple ultra-high-performance liquid chromatography–mass spectrometry (UHPLC–MS) methods and reference in-house library of authenticated metabolite standards. 455 metabolite annotations, including annotated degradation products, spanning 9 super-pathways and 85 sub-pathways were analyzed (Supplementary Data 1). Quantification was based on area-under-the-curve measurements, normalization to internal standards and to biological material as described below.[5] Nine invariant metabolites were removed, and missing data were imputed using one-fifth of minimum positive values.

*Targeted metabolomics data collection*

For each sample, 20 µL CSF or 5 µL plasma was extracted by brief sonication in 200 µL or 300 µL extraction buffer, respectively (80% methanol, 25 mM ammonium acetate, 2.5 mM sodium ascorbate in LC–MS–grade water), supplemented with isotopically labeled amino acids (Cambridge Isotope Laboratories, MSK-A2-1.2, Tewksbury, MA USA), aminopterin, and reduced glutathione (CNLM-6245-10). After centrifugation (10 min, maximum speed), supernatants were dried under nitrogen and reconstituted in 17 µL (CSF) or 25 µL (plasma) LC–MS–grade water containing QReSS (MSK-QRESS-KIT), vortexed, centrifuged, and transferred to LC–MS vials. One microliter was injected for analysis. A pooled quality-control (QC) sample was generated and diluted 3- and 10-fold to monitor analytical stability and signal linearity. Chromatographic separation was performed on a ZIC-pHILIC column (150 × 2.1 mm, 5 µm; EMD Millipore) using a Vanquish™ Flex UHPLC (Thermo Fisher Scientific, Waltham, MA, USA) with buffer A (acetonitrile) and buffer B (20 mM ammonium carbonate, 0.1% ammonium hydroxide, pH ~9). The gradient was 20–80% B over 20 min, returned to 20% B at 20.5 min, and held until 28 min at 150 µL/min; column and autosampler temperatures were 25 °C and 4 °C, respectively. Mass spectrometry was performed on a Q Exactive Orbitrap (Thermo Fisher Scientific) with an Ion Max source and HESI II probe in positive and negative ionization modes (m/z 70–1000). Settings were: resolution 70,000; AGC target $1 \times 10^6$; maximum injection time 20 ms; sheath/auxiliary/sweep gas 35/8/1; spray voltage 3.5 kV (positive) or 2.8 kV (negative); capillary temperature 320 °C; S-lens RF 50; auxiliary gas heater 350 °C. Metabolites assessed by targeted metabolomics are listed in Supplementary Data 2.

*Data processing and normalization for semi-targeted and targeted metabolomics*

For targeted metabolomics analysis, relative quantitation of polar metabolites was performed using a combination of TraceFinder version 5.1 (Thermo Fisher Scientific, Waltham, MA) and Compound Discoverer (CD) version 3.3 to increase annotation efficiency and data completeness. Both platforms referenced in-house retention time libraries, including a curated panel of ~500 compounds and 37 isotopically labeled internal standards (**Supplementary Data 2**). For TraceFinder analysis, metabolites were matched within a 5 ppm mass tolerance against the in-house library. For CD analysis, positive and negative ionization modes were processed separately. Filtering steps were based on signal-to-noise thresholds, ppm error, formula annotation, and comparison to blank injections, with features retained if abundance was >3-fold higher in true samples. Retention time correction was applied with a tolerance of 40 seconds, reflecting the stability of the HILIC method. Where metabolites were detected in both ionization modes, the value from the mode with higher integrated intensity was retained.

The output from both platforms was merged into a single metabolite area list for downstream quality control and normalization and was processed with an in-house R script:

(https://github.com/FrozenGas/KanarekLabTraceFinderRScripts/blob/main/MS_data_script_v2.4_20221018.R).

Pooled samples and fractional dilutions were prepared as quality controls, and only those metabolites were taken forward for analysis for which the coefficient of determination ($R^2$)

between dilution factor and peak area was ≥0.8 (high-confidence metabolites) and for which the coefficient of variation (CV) was below 30%. Normalization for biological material amounts was based on the total integrated peak area values of high-confidence metabolites within an experimental batch after normalizing to the averaged factor from all mean-centered chromatographic peak areas of isotopically labeled amino acids internal standards.

Semi-targeted raw data from the Metabolon platform was similarly processed with our in-house script, performing corresponding normalization for biological material based on the total integrated peak area values of high-confidence metabolites. Further data scaling (Pareto) and transformation ($Log_{10}$) were performed within the MetaboAnalyst platform version 5.0-6.0.[6]

*Differential statistics analysis for semi-targeted and targeted metabolomics*

For the exploratory differential and pathway analyses shown in Figs 2 and 3, data from each platform were processed within MetaboAnalyst.[6] Metabolites with >50% missing values were excluded and remaining missing values were imputed based on the limit of detection; for the semi-targeted dataset an interquartile range filter of 10% was applied (not applied for the urinary outcome). Data were log-transformed and Pareto scaled. Volcano-plot p-values were adjusted using the Benjamini–Hochberg false discovery rate (FDR). Pathway analysis used the KEGG *Homo sapiens* library[7] with metabolite names mapped to HMDB identifiers; uncertain or isomeric annotations were mapped to the first listed compound, and automatic name matches were checked by hand. PLS-DA was assessed by 5-fold cross-validation ($R^2$, $Q^2$, accuracy) and a permutation test, and variable importance by VIP scores (Supplementary Fig. 4B–J). No sample was excluded on the basis of principal component analysis. Fold changes are expressed as improved/not improved throughout; values above 1 indicate higher levels in patients who improved.

For the biofluid comparisons (Fig. 6 and Supplementary Fig. 8), group differences were tested by one-way ANOVA with Šídák's multiple comparisons test or t-tests in Prism version 11.0.0 (GraphPad, Boston, MA, USA). Fold changes and stars on the pathway maps (Fig. 5 and Supplementary Fig. 2C–D) come from the adjusted per-metabolite models described below, so that one test is used throughout.

*Correlation analysis between targeted and semi-targeted metabolomics datasets*

Metabolites detected in both semi-targeted and targeted LC–MS datasets were matched based on metabolite identity and analyzed across overlapping samples. Data were processed in R Studio (version 2025.05.1+513) using the tidyverse package. For each metabolite, Pearson correlation coefficients were calculated using log10-transformed and Pareto-scaled values to assess concordance in relative variation across samples. Pareto scaling was performed by mean-centering and dividing by the square root of the standard deviation. Agreement in absolute measurements was evaluated using Bland–Altman analysis[8] on raw concentration values. For each metabolite, the log10-transformed difference between platforms was calculated, and the mean difference (bias) and 95% limits of agreement (LoA; mean $\pm$ 1.96 $\times$ SD) were derived. The width of the LoA interval (expressed as fold difference) was used as a measure of quantitative agreement. Correlation coefficients and LoA widths were combined into a two-dimensional representation for visualization. Metabolites belonging to significant modules were annotated in the plot.

*Locked dataset and statistical analysis plan*

Before any association with outcome was examined, the analysis dataset was locked: cohort membership followed the clinical team's final cohort table, outcomes were coded per domain at 3 months (worsened pooled with not improved; worsening retained as a separate flag; patients without a 3-month assessment of a domain were omitted from the analyses of that outcome, and no outcome was imputed), and sample exclusions were technical only, defined by a quality-control rule fixed in advance (median of glutamine, creatinine, citrate and alanine below 10% of typical ventricular CSF, or more than 20 missing metabolites). One targeted ventricular sample was not acquired and one failed this rule; no sample was removed on the basis of its metabolite profile. The statistical analysis plan, including the indices, models, covariates and multiple testing families, was fixed on 12 October 2026, before the outcome associations reported here were run. The pathways summarized by the indices came from the semi-targeted discovery analysis, and the choice of their component metabolites was also informed by an initial exploratory analysis of the targeted data; the index tests are therefore prespecified with respect to this analysis but not independent of the data that motivated them. Every subsequent change (one patient's inclusion following the clinicians' cohort table, one corrected semi-targeted value, and the analyses added later) is dated in an analysis log released with the code; the patient inclusion and the corrected value did not change any outcome result.

*Prespecified indices and primary analysis*

Two composite indices and eleven ratios summarizing the discovery modules were defined, with component signs fixed from pathway position, and computed outcome-blind. The one-carbon/redox index (P1) is the mean of signed z-scores of log2 cystathionine (+), 2-hydroxybutyrate (−), 2-aminobutyrate (−), 5-methyltetrahydrofolate (+), phosphocholine (+) and phosphoethanolamine (+); the kynurenine index (P2) is the mean z-score of log2 tryptophan, kynurenine and their ratio. Ratios were log2(numerator) − log2(denominator): kynurenine/tryptophan, kynurenic acid/kynurenine, quinolinic acid/kynurenic acid, 5-HIAA/tryptophan, cystathionine/methionine, 2-hydroxybutyrate/cystathionine, 2-aminobutyrate/2-hydroxybutyrate, phosphocholine/choline, α-ketoisovalerate/valine, lactate/pyruvate, and (butanoate + pentanoate)/hexose. Non-detects were set to half the lowest detected value; a composite was missing if more than half its components were imputed. The primary outcome was the number of domains improved (0–3) among domains symptomatic at baseline, analyzed by proportional-odds regression on the index (per SD) adjusted for age, sex and enrollment cohort (n = 43), with likelihood-ratio p-values and profile-likelihood 95% confidence intervals.[9] The analysis plan did not name the test; Wald tests were used first and were replaced by likelihood-ratio tests after review by the study statistician, because Wald tests understate significance for large effects in small samples (Wald results in the released code). P1 and P2 formed the primary family and the eleven ratios the secondary family, each with Benjamini–Hochberg correction.[10] Per-domain logistic models were exploratory. Because ordinary maximum likelihood showed quasi-complete separation for two ratios in the cognition model (8 non-improved patients), all three domains were fitted by Firth's penalized likelihood, with intervals and p-values from the penalized profile likelihood.[11] Sensitivity analyses for all indices (Supplementary Table 1b): (a) excluding patients who worsened in any domain, (b) iNPH only, (c) excluding three samples with flagged normalization factors, (d) cohort 2 only, (e) adding body mass index, (f) patients with all three domains scored, (g) a quasi-binomial model of the number of domains improved out of the number scored, and (h) excluding index values with |robust z| > 3.5; in (a), (b), (d) and (f), outcome categories with fewer than five patients were merged with the adjacent category. An earlier sensitivity analysis excluding the patient with the most extreme cystathionine value is superseded by (h). Odds ratios above 1 indicate that higher index values accompany more improvement.

*Exploratory per-metabolite analysis*

Every endogenous targeted metabolite detected in at least half of the samples (129 metabolites) was tested with the same models as the indices, per domain and for the number of domains improved, after $\log_2$ transformation and z-scoring, with Benjamini–Hochberg correction within each outcome. Non-endogenous metabolites (microbial, dietary, drug-derived or of uncertain origin; 27 metabolites) formed a separate family. The same models were applied to semi-targeted metabolites (without the cohort covariate, which is constant in cohort 1) for the pathway maps and to metabolites previously reported in NPH (Table 2). These analyses are exploratory.

*Prediction models*

Prediction followed a locked protocol (Supplementary Fig. 4A). Each outcome was modelled from the 156 targeted metabolite columns of the 55 ventricular samples (patients with a missing outcome were omitted: cognition n = 38, gait n = 43, urinary n = 38). Performance was estimated by nested cross-validation[12]: outer stratified 5-fold cross-validation repeated 20 times (100 test folds, identical for every model), with an inner stratified 5-fold loop for all tuning and feature selection. Folds were stratified by outcome; no class reweighting or resampling was applied in the locked protocol. Preprocessing ($\log_{10}$ transformation, removal of metabolites with >20% missing values in the training fold, median imputation, standardization) was fitted on each training fold only. The prespecified primary model was L2-penalized (ridge) logistic regression on all metabolites, tuned by inner log-loss over 11 log-spaced penalty values ($C = 10^{-3}$ to $10^{2}$). Comparators were a linear SVM and an RBF-kernel SVM, each trained on the top-k metabolites ranked by the absolute coefficients of a logistic regression fitted in the training fold ($k \in \{5, 10, 20, 30, 50\}$; $C \in \{0.01, 0.1, 1, 10\}$; tuned by inner AUC). The RBF-kernel SVM corresponds to the model of our earlier analysis, which used scikit-learn's default kernel. Out-of-fold predictions were pooled per repeat; we report the median AUC across repeats with its range, and for the ridge model the Brier score, calibration slope and intercept,[13] and classification metrics at a threshold fixed in advance (predicted probability of improvement below the cohort prevalence; SVM decision score below zero). Significance was assessed by permuting outcome labels 1,000 times and rerunning the whole nested pipeline (5 outer repeats per permutation), with p = (1 + number of null AUCs ≥ observed)/(1 + 1,000) and a max-over-models correction for the choice among three models. Incremental value was assessed on the same folds by comparing ridge models with clinical covariates (age, sex, cohort), clinical covariates plus P1 and P2 recomputed within each training fold, and clinical covariates plus the full metabolome; log-loss was the primary comparison because the clinical-only model shrank to near-constant predictions. Adding the short-chain fatty acid/hexose ratio was an exploratory analysis added after inspection of the results. Model fairness and differences in performance between sociodemographic groups were not assessed, given the sample size. Analyses used Python 3.13 and scikit-learn 1.9.1.[14] For comparison, the pipeline of our initial exploratory analysis (leave-one-out cross-validation repeated over 10 random seeds, with the number of metabolites selected on the same folds used for evaluation) was re-run on the locked cognition data together with a permutation null of that tune-and-report procedure (100 permutations); these estimates and the originally reported values are given in Supplementary Table 3d.

*Nested leave-one-out re-analysis of the initial pipeline*

After the analysis lock, the leave-one-out pipeline of the initial analysis was re-run in nested form, first for cognition by the study statistician and then re-implemented in the released code (step 40) for all three outcomes, which gave the estimates reported here (Fig. 4B, D, F; Supplementary Fig. 6; Supplementary Table 3d). Preprocessing followed the initial analysis: values were log2-transformed and, within each training fold, metabolites with more than 20% missing values were removed, remaining missing values were replaced by the training-fold mean and metabolites were autoscaled (training-fold mean and SD). Within each training fold, metabolites were then ranked by the absolute coefficient of an L2-regularized logistic regression with balanced class weights, and the top k were used to fit a linear SVM with balanced class weights and regularization parameter C. $k \in \{10, 15, 20, 25, 30\}$ and $C \in \{0.01, 0.1, 1, 10\}$ were chosen jointly by the AUC of a stratified 5-fold cross-validation inside the training fold (with preprocessing refitted in each inner fold), so that the held-out patient's prediction was not used to select either hyperparameter. This nested version differs from the initial analysis in three respects: the kernel is linear, as described in the original report (the initial code used scikit-learn's default RBF kernel), class weights are balanced, and k and C are tuned inside the training fold instead of being fixed at the values with the best leave-one-out AUC ($k = 30$, $C = 10$). The held-out decision values, each from the model fitted without that patient, were concatenated and a single AUC computed against the observed outcomes. The procedure was repeated with 10 random seeds, which change only the inner split. The mean AUC across seeds and its range are reported, with a 95% confidence interval from 5,000 bootstrap resamples of patients (AUC recomputed per resample and averaged over seeds). Significance was assessed by 1,000 label permutations of the complete pipeline for each outcome (one seed; the same number as for the locked protocol; permutation sequences fixed per outcome), $p = (1 + \text{number of permuted AUCs} \geq \text{observed mean}) / 1{,}001$, uncorrected for the choice among models. Feature-selection frequency was tallied over the outer folds of all seeds. Cognition ($n = 38$, 8 not improved): 0.79 (0.63–0.90; 95% CI 0.65–0.92), null mean 0.48, 95th percentile 0.75, $p = 0.028$. Gait ($n = 43$, 14 not improved): 0.65 (0.57–0.78; 95% CI 0.51–0.79), null mean 0.48, 95th percentile 0.71, $p = 0.12$. Urinary ($n = 38$, 13 not improved): 0.63 (0.56–0.72; 95% CI 0.49–0.77), null mean 0.48, 95th percentile 0.73, $p = 0.16$. The statistician's implementation used natural-log-transformed values with values below the limit of detection set to zero and gave a mean AUC of 0.83 for cognition (range 0.73–0.90; 95% CI 0.69–0.94; 200 permutations, $p = 0.030$); a first version of the released code without log transformation also gave 0.83 (Supplementary Table 3d).

*Transcriptomic integration*

Transcriptomic data were the previously reported RNA sequencing of CSF extracellular vesicles from cohort 1 (variance-stabilized counts; 14,294 genes detected in at least half of the samples)[3]; 32 patients had a cognitive outcome and 31 also had targeted metabolomics. Nineteen enzyme–

metabolite pairs with expected directions were fixed and committed before the transcriptomic matrix was opened and tested by one-sided Spearman correlation with Benjamini–Hochberg correction; pairs whose genes were detected in fewer than half of the samples were not tested. For the pathway-level analysis, KEGG gene sets[7]; one-carbon pool by folate as the primary set; cysteine and methionine, glutathione, glycerophospholipid, tryptophan and branched-chain amino acid degradation as secondary metabolic sets; cell cycle as a non-metabolic control) were fixed before analysis. For each gene, the outcome t-statistic was computed from a linear model adjusted for age and sex; the set statistic was the mean t of the set minus the mean t of all detected genes (a competitive test[15]), and significance was assessed by permuting patient outcome labels 1,000 times (seed fixed in advance), with Benjamini–Hochberg correction across the six metabolic sets. Per-patient set scores were correlated with P1 as a cross-layer check. This pathway-level test was defined after the enzyme–metabolite pairs had been found null and replaces a likelihood-ratio test of a gene model extended with metabolites selected after inspection of the data, which we did not retain because selection on the same data inflates it.

*Biofluid concordance and sample size*

Ventricular–lumbar concordance of indices and metabolites was assessed outcome-blind in the 12 matched patients by Spearman correlation, intraclass and concordance correlation, Bland–Altman analysis and two one-sided tests[16] for equivalence within a ±1.5-fold margin fixed in advance; lumbar indices were computed with the ventricular means, standard deviations and half-minimum values. Plasma values were multiplied by 8.8 to account for the lower injected volume-equivalent. Precision and validation sample size were derived from the locked group sizes using the Hanley–McNeil standard error of the AUC (Supplementary Table 3e–f).[17]

*Precision and validation sample size*

The standard error of the AUC was computed with the Hanley–McNeil formula,[17] which depends on the AUC and the numbers of improved and non-improved patients. For the locked group sizes, the attainable 95% confidence half-width (1.96 × SE) is reported for each outcome in Supplementary Table 3e; for cognition (8 non-improved of 38) it is ±0.21 at an AUC of 0.75. The validation sample size was defined as the smallest N at which the half-width for a true AUC of 0.75 reaches ±0.075. Because the standard error is dominated by the smaller outcome group, the cohort was sized for the outcome with the lowest non-improvement rate, cognition (8/38, 21%), yielding N = 293. In a cohort of this size, the gait and urinary outcomes (14/43, 33%; 13/38, 34%) would be estimated with half-widths of ±0.063 and ±0.062, respectively; at a 25% non-improvement rate, N = 254 would meet the same criterion (Supplementary Table 3f). DeLong intervals in simulated cohorts[1] (binormal scores with an AUC of 0.75, 500 cohorts per size) were slightly narrower than the Hanley–McNeil intervals, so these sizes are conservative (Supplementary Fig. 9).

**Supplementary Data**

**Supplementary Data 1 lists all 455 metabolite annotations (including annotated degradation products)** measured on the semi-targeted UHPLC–MS platform (Metabolon) after quality control. Each annotation is assigned with its super-pathway (9 in total) and sub-pathway (85 in total) and with HMDB, KEGG and PubChem identifiers.

**Supplementary Data 2** lists the metabolites on the targeted LC–MS platform: 155 distinct metabolites passing quality control, referenced against the in-house library of about 500 authenticated compounds and 37 isotopically labelled internal standards. The file gives annotation and detection rates and flags known duplicate integrations.

**Clinical metadata and 3-month outcomes for all patients in the metabolomics analyses can be made available upon request (see Data availability)** (n = 86):

- semi-targeted ventricular CSF, n = 42
- targeted ventricular CSF, n = 55 (2 technical exclusions)
- targeted lumbar CSF, n = 29
- targeted plasma, n = 33

31 patients have ventricular CSF in both the semi-targeted and the targeted datasets.

**Supplementary Table 1.** Association of prespecified metabolic indices with outcome:

- (a) primary analysis: number of domains improved (proportional odds) and cognitive improvement (Firth logistic regression), with FDR within each prespecified family
- (b) sensitivity analyses
- (c) per-domain results for cognition, gait and urinary continence

**Supplementary Table 2.** Exploratory per-metabolite scan of targeted ventricular CSF: covariate-adjusted associations of each endogenous metabolite with cognitive improvement and with the number of domains improved, with FDR.

**Supplementary Table 3. Outcome prediction by nested cross-validation and study precision:**

- (a) performance metrics for ridge, linear SVM and nonlinear SVM (RBF kernel) by outcome
- (b) permutation tests, with max-T correction across models
- (c) added value of metabolic indices over clinical predictors (log-loss ladder, including the post hoc R11 step)
- (d) estimates from the initial exploratory pipeline as originally reported, with the 2026 audit re-run
- (e) detectable effect sizes and AUC precision for the locked group sizes; (f) validation sample size (Hanley–McNeil standard error of the AUC)

**Supplementary Table 4.** Integration with CSF extracellular-vesicle transcriptomics (cohort 1):

- (a) prespecified gene–metabolite pairs, targeted platform (primary)
- (b) the same pairs on the semi-targeted platform (replication)
- (c) KEGG gene-set competitive tests (one-carbon pool by folate as the primary set) for cognitive improvement
- (d) the same gene-set tests for the number of domains improved
- (e) cross-layer correlations of gene-set scores with the one-carbon/redox index

**Supplementary Table 5.** Agreement between biofluids: ventricular vs lumbar CSF and plasma for (a) the metabolic indices and (b) individual metabolites.
